# Environmental impacts associated with UPF consumption: which food chain stages matter the most? *Findings from a representative sample of French adults*

**DOI:** 10.1101/2022.05.28.22275717

**Authors:** Emmanuelle Kesse-Guyot, Benjamin Allès, Joséphine Brunin, Hélène Fouillet, Alison Dussiot, Florine Berthy, Elie Perraud, Serge Hercberg, Chantal Julia, François Mariotti, Mélanie Deschasaux-Tanguy, Bernard Srour, Denis Lairon, Philippe Pointereau, Julia Baudry, Mathilde Touvier

## Abstract

**Background:** Scientific literature about environmental pressures associated with dietary patterns has been considerably growing over the last decade. However, few studies have analyzed the environmental impacts associated with the consumption of ultra-processed food (UPF) and which steps of the food system that contribute most to environmental pressures. The objective of this study was to investigate, in a representative sample of the adult French population, the environmental pressures of diets according to UPF consumption.

**Methods:** The study was conducted in 2,121 adults of the French nationally representative survey INCA 3. Food intakes were analyzed to define the %UPF (in weight) in the diet according to NOVA classification. Using detailed environmental data of foods of Agribalyse, we could assess the contribution of UPF to 14 indicators of environmental pressure and details those related to the stage of the food consumed: production, processing, storage, packaging, transport and retailing at the food level. The data was described according to quintiles of % of UPF in the diet and analyzed using crude and energy-adjusted models.

**Results:** Compared to low consumers of UPF (Q1, median UPF= 7%), high consumers (Q5, median UPF= 35%) had a higher energy intake (+22%) which came along with different environmental pressures (e.g. +15% greenhouse gas emissions (GHGe), +17% land use, -7% water use and +8% cumulative energy demand). Higher pressures could be ascribed to higher energy. However, the processing and packaging stages were significant contributor to energy demand. In Q5, contributions of the UPF category to total pressure were 35%, 39%, 28% and 42% for GHGe, water use land use, and energy demand, respectively, while ranging from 11 to 15% in Q1.

**Conclusions:** Diets rich in UPF, compared to low, were overall associated with intensification in GHGe, land use, and energy demand and with higher contribution of post-farm stages, in particular processing regarding energy demand.

**Highlights:**

- Overall, higher UPF consumption was associated with higher environmental impact, in particular GHGE and land use
- Low UPF consumers had overall higher water footprint (due to their high fruit and vegetable intake)
- A large part of the higher pressures observed among participants with higher UPF consumption was explained by their higher dietary energy intake since the associations did not remain or were even reversed after energy adjustment
- Regardless of the % UPF in the diet, GHGe, land use and water use pressures mostly occurred at the stage of agricultural production, in contrast, packaging and processing stages were also important contributors to energy demand
- Contribution of the UPF category to total impact/pressure varied across indicators, with a high contribution of the UPF category to energy demand, due to the packaging and processing stages, but a low contribution to GHGe and land use, because higher consumers of UPF are lower consumers of animal products

## Introduction

In the recent decades, food systems and food supplies have become largely globalized with changes occurring at all steps of the food chain, from production to consumption (1). Urbanization and modernization have profoundly changed eating habits (2). Diets first in Western industrialized countries and now in many lower and middle-income countries are characterized by high consumption of animal products but also salt, fat and sugar, and recognized as major risk factors for many chronic diseases (3). Besides, the consumption of “ultra-processed foods” (UPF), has grown around the world and now reaches more than half of daily energy intake in the UK and US (4) and is 31% in France (5). These are foods that underwent extensive chemical or physical transformations and/or containing cosmetic food additives or other industrials ingredients (e.g. hydrogenated oils, fructose syrup, etc.). Although not systematically, they often contain on average higher amounts of saturated fat, salt, and sugar and lower amounts of fibers, micronutrients and potentially healthy active compounds (6). In the last decade, a growing body of studies suggesting a role of UPF consumption on health has emerged. This literature is broadly growing, leading to the first reviews and meta-analyses summarizing findings from prospective studies and consistently showing associations between UPF consumption and increased risk of many non-communicable diseases (4,7–13). Most of the studies are based on the NOVA classification distinguishing unprocessed, minimally processed, processed and ultra-processed foods (6).

Beyond health issues, the current global food system greatly contributes to the degradation of the environment by undermining natural resources, including water and forests, and jeopardizing climate stability by increase of greenhouse gas emissions (GHGe) (14,15). However, even if the raw material production stage is the most impacting for the environment, post-production steps (e.g. processing, packaging, and transport) in the food industry are also resource-intensive (16,17) and are important contributors of GHGe and energy demand. A recent time-series study conducted in Brazil showed that the share of unprocessed foods in the diet decreased over a 30-year period, while the share of processed and UPF increased, especially UPF based on animal products, doubling their contribution to total diet-related environmental impacts over the same period (18).

The shortcomings of studies investigating environmental pressures associated with UPF have been described in the literature and include, among other things, the failure to take into account some ingredients and imprecise data on the processing and packaging stages (19). In addition, scientific literature documenting environmental pressures related to UPF intakes is very scarce (20) and focused on data from life cycle assessment (LCA) covering the entire chain, without individualizing its different stages (production, processing, storage, packaging, transport and retailing). However, some authors reported that consumption of discretionary foods (foods high in saturated fats, sugars, salt and/or alcohol that can be eaten occasionally in small amounts, but are not a necessary part of the diet) may contribute to an important part of diet-related environmental pressures (including water use, energy use and GHGe). Energy density of such eating habits could be a strong determinant of environmental pressures (21,22). In addition, the food processing sector is an important contributor to total food loss and waste (along the entire food supply chain) generated by retailers and consumers (23).

While the role of UPF on human health, based on the NOVA classification, is becoming well documented, it is now important to estimate, using a systematic methodological approach, the impact of UPF on planetary health, as part of a holistic approach to health.

In this context, the aim of the present study was to estimate environmental pressures of UPF consumption, 1) overall, 2) adjusted for energy intake to account for level of consumption, 3) differentiating the different stages of the whole food chain, and 4) by NOVA group, in a French representative study that included a large set of environmental indicators.

## Methods

### Population

This study was based on the French nationally representative survey INCA 3 conducted in 2014-2015 by the French Agency for Food, Environmental and Occupational Health & Safety (ANSES) and including 2,121 adult participants who provided valid dietary consumption data (24). The design of the study as well as the methods have been detailed elsewhere (24).

Participants were selected according to a three-stage random sampling plan (geographical units, dwellings then individuals) drawn at random by the National Institute of Statistics and Economic Studies (INSEE), based on the annual population census in 2011. One individual per dwelling was then drawn at random from among the eligible individuals at the time of contact with the household. Individual weight was calculated according to INSEE method to improve representativeness according to region, size of the urban area, occupation and socio-professional category of the household reference person, size of the household, level of education, sex and age (25). The INCA 3 study protocol was authorised by the National Commission on Informatics and Liberty, after a favourable opinion from the Advisory Committee on Information Processing in Health Research (CCTIRS). The study also received a favourable opinion from the Conseil National de l’Information Statistique (CNIS) on 15 June 2011 (n°121/D030) and was awarded the label of “general interest” and statistical quality by the INSEE Label Committee (n°47/Label/D120). The data collected in the INCA 3 study are available on the website https://www.data.gouv.fr/fr/datasets/donnees-de-consommations-et-habitudes-alimentaires-de-letude-inca-3/.

The data collected in the INCA 3 cross-sectional study included food and drink consumption and socio-demographic and lifestyle characteristics.

### Dietary data

Detailed consumption data were collected over 3 non-consecutive days (2 weekdays and 1 weekend day) distributed over approximately 3 weeks, using the 24-hour recall method conducted by telephone by trained interviewers using a standardised software (GloboDiet)(26). The quantification of portion sizes was carried out using a picture booklet of food portions and household measurements.

Dietary intakes were calculated using the French nutritional composition data from the 2016 food composition database published by the French Information Centre on Food Quality (27).

Mixed foods were decomposed using the standardized recipes validated by dieticians.

All food items were classified according to the NOVA classification (6,28) as previously extensively described (29). At the individual level, the percentages (in weight) of the diet in NOVA1 (unprocessed or minimally processed foods), NOVA2 (culinary ingredients), NOVA3 (processed foods), and NOVA4 (UPF) were computed as described in the **Supplemental Material 1**.

The overall quality of the diet was assessed using two dietary scores, namely the sPNNS-GS2 (30) and the PANDiet (31), which have extensively been described. Further details are presented in **Supplemental Material 2**.

### Environmental indicators

Diet-related environmental pressures were estimated using data from the French database Agribalyse® 3.0.1 developed by the French Agency for the Environment and Energy Management (ADEME). Agribalyse® 3.0.1 contains environmental indicators for 2,497 foods consumed in France for which nutritional contents is also available (32) using the same taxonomy. A total of 14 midpoint indicators were available: GHGe, ozone depletion, particulate matters, ionizing radiation (effect on human health), ecotoxicity, photochemical ozone formation (effect on human health), acidification, terrestrial eutrophication, freshwater eutrophication, marine eutrophication, land use, water use, resource use, minerals and metals and resource use, fossils and one endpoint ecological footprint (EF) calculated according to the product environmental footprint (PEF) methodology (33).

Environmental indicator estimations were based on the method of LCA whose scope is “from field to plate”. The perimeter of the indicators covers each step of the value chain: agricultural production, transport, processing, packaging, distribution and retailing, preparation at the consumer’s level and disposal of packaging. These different stages have been split into two phases 1) production and 2) post-farm. The methodology has been extensively explained in *ad hoc* published reports (34,35) summarized in **Supplemental Material 3**.

### Statistical analysis

Participants were ranked and divided into weighted quintiles of %UPF. Socio-demographic and dietary characteristics were described across weighted quintiles of %UPF using ANOVA, or ANCOVA models when adjustment for energy intake was performed. Micronutrient and fiber intakes were adjusted for energy intake using the residual method (36) and macronutrients were reported as % of total energy intake.

Main food group contribution to %UPF by quintiles were also described.

In the main analysis, diet-related environmental footprints, according to quintiles of %UPF in the diet, were first estimated overall, using crude and energy-adjusted ANOVA and ANCOVA models. For the 4 indicators that are well documented in the literature and therefore the most robust (GHGe, water use, land use, and energy demand), we also examined the contribution of the different food system stages to the environmental pressures. Finally, the respective contribution of the different NOVA food category to the different environmental pressures was assessed.

Several sensitivity analyses were conducted: 1) models were adjusted for %NOVA1 in the diet as NOVA1 consumption is inversely correlated to UPF, 2) UPF vs. %NOVA1 substitution was modeled by adjustment for energy intake, %NOVA2 and %NOVA3 (36) for each indicator and farm and post-farm steps, and 3) the main analysis described above was reperformed using the %UPF as % of total energy intake instead of total weight. For this purpose, due to distribution of individual weightings, quartiles were considered.

All tests were two-sided and a P-value <0.05 was considered significant. Statistical analyses were performed using SAS Software (version 9.4, SAS Institute Inc, Cary, NC, USA) and figures were performed developed using R version 3.6.

## Results

The characteristics of the total study population and according to quintiles of %UPF are presented in **Table 1**. The studied population included 2,121 participants (58% women), with a mean age of 47 years (SD=16). In this population, %UPF was 18.16 (SD=11.66) and 20.59 (SD=12.06) in women and men, respectively. Participants with higher %UPF were more often male, younger, less educated, with lower income and were more often unemployed or students. In addition, %UPF was inversely related to proportion of NOVA1 in the diet (Q5 vs. Q1=-38%).

**Table 1:**
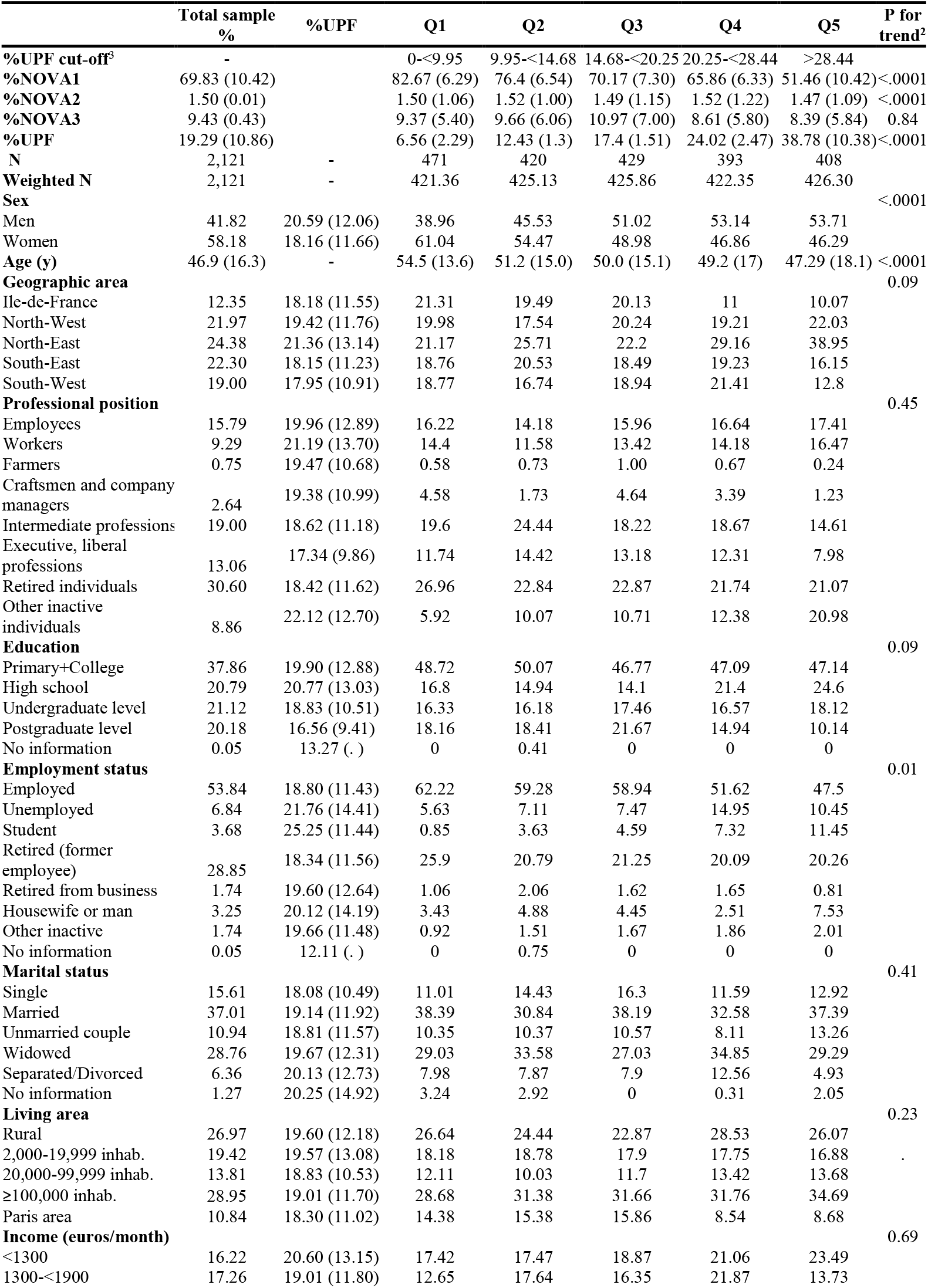
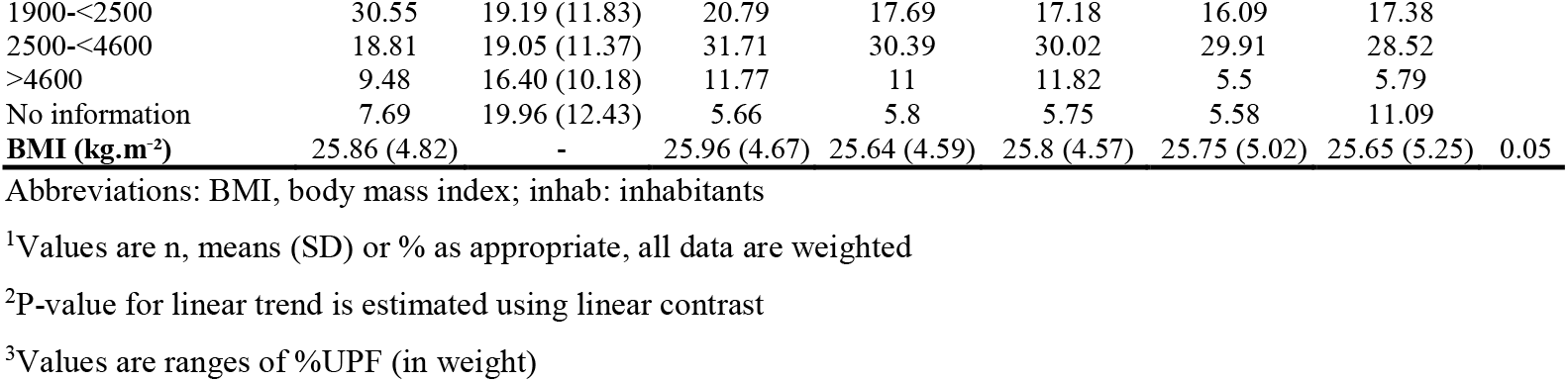
Characteristics of study participants according to quintiles of %UPF, (INCA 3, n=2,121)^1^.

Dietary consumption greatly differed according to the %UPF in the diet (**Table 2 and Supplemental Table 1**). In energy-adjusted models (**Table 2**), participants with high %UPF (Q5), compared to low (Q1), had higher consumption of sweetened beverages, fruit juices, legumes, soup, and prepared dishes, and lower consumption of non-sweetened beverages, whole grain and refined cereals, fruits, vegetables, animal and vegetable fat, eggs, fish, and red meat. No linear relationship was observed for alcoholic beverages, sweet and fat foods, condiments, potatoes, dairy products, processed meat, and poultry.

**Table 2:**
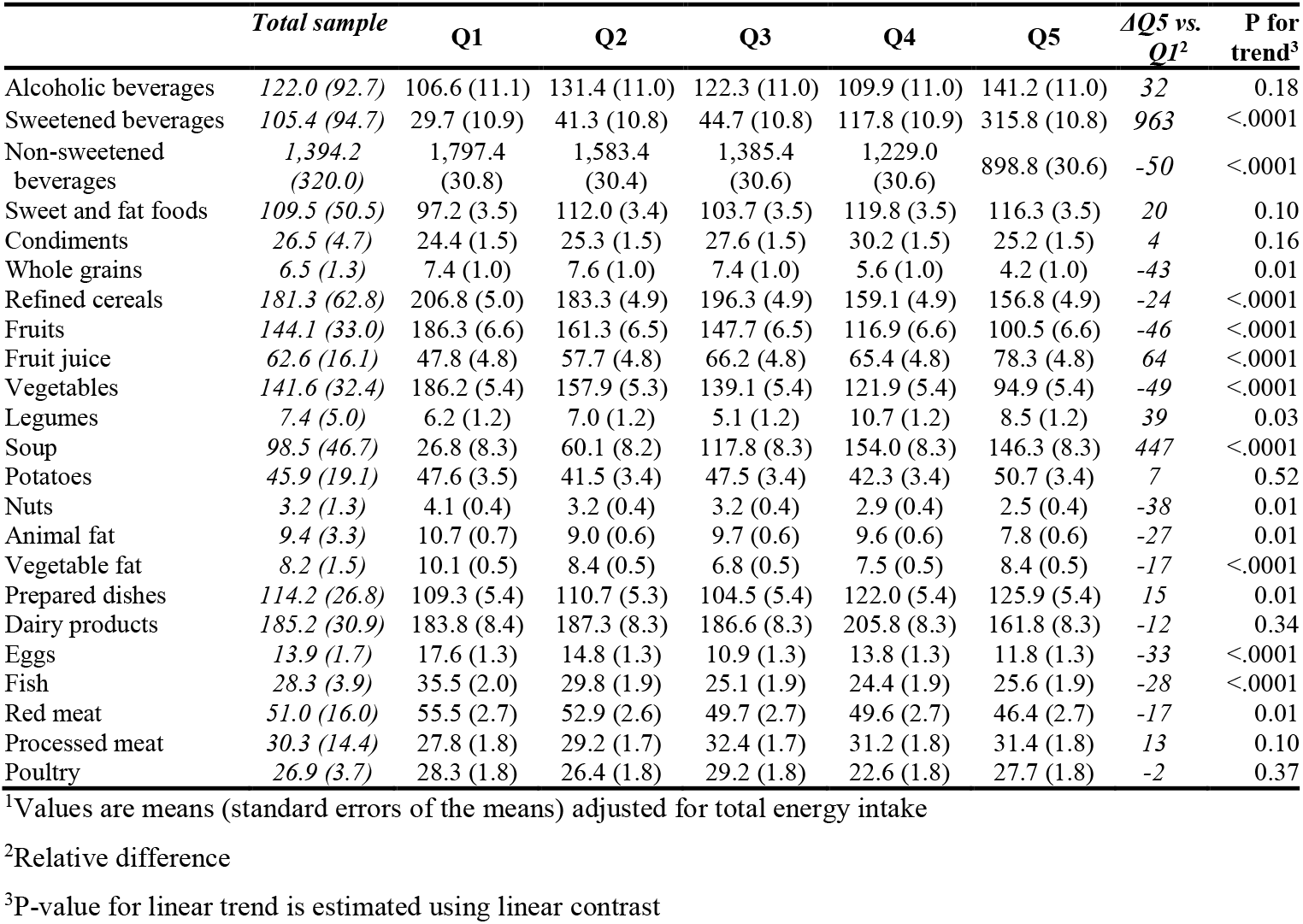
Consumption of food groups (g/d) according to %UPF quintiles, (INCA 3, N=2,121)^1^.

|  | *Total sample* | **Q1** | **Q2** | **Q3** | **Q4** | **Q5** | **$\Delta Q5$ vs.**<br>**$Q1^2$** | **P for trend<sup>3</sup>** |
| --- | --- | --- | --- | --- | --- | --- | --- | --- |
| Alcoholic beverages | 122.0 (92.7) | 106.6 (11.1) | 131.4 (11.0) | 122.3 (11.0) | 109.9 (11.0) | 141.2 (11.0) | 32 | 0.18 |
| Sweetened beverages | 105.4 (94.7) | 29.7 (10.9) | 41.3 (10.8) | 44.7 (10.8) | 117.8 (10.9) | 315.8 (10.8) | 963 | <.0001 |
| Non-sweetened beverages | 1,394.2 (320.0) | 1,797.4 (30.8) | 1,583.4 (30.4) | 1,385.4 (30.6) | 1,229.0 (30.6) | 898.8 (30.6) | -50 | <.0001 |
| Sweet and fat foods | 109.5 (50.5) | 97.2 (3.5) | 112.0 (3.4) | 103.7 (3.5) | 119.8 (3.5) | 116.3 (3.5) | 20 | 0.10 |
| Condiments | 26.5 (4.7) | 24.4 (1.5) | 25.3 (1.5) | 27.6 (1.5) | 30.2 (1.5) | 25.2 (1.5) | 4 | 0.16 |
| Whole grains | 6.5 (1.3) | 7.4 (1.0) | 7.6 (1.0) | 7.4 (1.0) | 5.6 (1.0) | 4.2 (1.0) | -43 | 0.01 |
| Refined cereals | 181.3 (62.8) | 206.8 (5.0) | 183.3 (4.9) | 196.3 (4.9) | 159.1 (4.9) | 156.8 (4.9) | -24 | <.0001 |
| Fruits | 144.1 (33.0) | 186.3 (6.6) | 161.3 (6.5) | 147.7 (6.5) | 116.9 (6.6) | 100.5 (6.6) | -46 | <.0001 |
| Fruit juice | 62.6 (16.1) | 47.8 (4.8) | 57.7 (4.8) | 66.2 (4.8) | 65.4 (4.8) | 78.3 (4.8) | 64 | <.0001 |
| Vegetables | 141.6 (32.4) | 186.2 (5.4) | 157.9 (5.3) | 139.1 (5.4) | 121.9 (5.4) | 94.9 (5.4) | -49 | <.0001 |
| Legumes | 7.4 (5.0) | 6.2 (1.2) | 7.0 (1.2) | 5.1 (1.2) | 10.7 (1.2) | 8.5 (1.2) | 39 | 0.03 |
| Soup | 98.5 (46.7) | 26.8 (8.3) | 60.1 (8.2) | 117.8 (8.3) | 154.0 (8.3) | 146.3 (8.3) | 447 | <.0001 |
| Potatoes | 45.9 (19.1) | 47.6 (3.5) | 41.5 (3.4) | 47.5 (3.4) | 42.3 (3.4) | 50.7 (3.4) | 7 | 0.52 |
| Nuts | 3.2 (1.3) | 4.1 (0.4) | 3.2 (0.4) | 3.2 (0.4) | 2.9 (0.4) | 2.5 (0.4) | -38 | 0.01 |
| Animal fat | 9.4 (3.3) | 10.7 (0.7) | 9.0 (0.6) | 9.7 (0.6) | 9.6 (0.6) | 7.8 (0.6) | -27 | 0.01 |
| Vegetable fat | 8.2 (1.5) | 10.1 (0.5) | 8.4 (0.5) | 6.8 (0.5) | 7.5 (0.5) | 8.4 (0.5) | -17 | <.0001 |
| Prepared dishes | 114.2 (26.8) | 109.3 (5.4) | 110.7 (5.3) | 104.5 (5.4) | 122.0 (5.4) | 125.9 (5.4) | 15 | 0.01 |
| Dairy products | 185.2 (30.9) | 183.8 (8.4) | 187.3 (8.3) | 186.6 (8.3) | 205.8 (8.3) | 161.8 (8.3) | -12 | 0.34 |
| Eggs | 13.9 (1.7) | 17.6 (1.3) | 14.8 (1.3) | 10.9 (1.3) | 13.8 (1.3) | 11.8 (1.3) | -33 | <.0001 |
| Fish | 28.3 (3.9) | 35.5 (2.0) | 29.8 (1.9) | 25.1 (1.9) | 24.4 (1.9) | 25.6 (1.9) | -28 | <.0001 |
| Red meat | 51.0 (16.0) | 55.5 (2.7) | 52.9 (2.6) | 49.7 (2.7) | 49.6 (2.7) | 46.4 (2.7) | -17 | 0.01 |
| Processed meat | 30.3 (14.4) | 27.8 (1.8) | 29.2 (1.7) | 32.4 (1.7) | 31.2 (1.8) | 31.4 (1.8) | 13 | 0.10 |
| Poultry | 26.9 (3.7) | 28.3 (1.8) | 26.4 (1.8) | 29.2 (1.8) | 22.6 (1.8) | 27.7 (1.8) | -2 | 0.37 |

<sup>1</sup>Values are means (standard errors of the means) adjusted for total energy intake<sup>2</sup>Relative difference<sup>3</sup>P-value for linear trend is estimated using linear contrast

Energy intake increased across quintiles (Q5 vs. Q1=+22%) (**Table 3**). Higher %UPF was associated with higher intake of carbohydrates and sugar, and lower intake of protein, fibers and most micronutrients (total and non-heme iron, copper, magnesium, potassium, selenium, vitamins C and B12, and zinc). Of note, for saturated fatty acids and lipids no linear trends were observed.

**Table 3:** Nutrient intakes and dietary scores according to %UPF quintiles, (INCA 3, n=2,121)^1^.

|  | Q1 | Q2 | Q3 | Q4 | Q5 | P for trend<sup>4</sup> |
| --- | --- | --- | --- | --- | --- | --- |
| Alcohol-free energy intake (kcal/d)<sup>2</sup> | 1,782.70 (34.81) | 1,949.69 (34.66) | 2,133.58 (34.63) | 2,110.19 (34.77) | 2,178.44 (34.61) | <.0001 |
| Lipids<sup>3</sup> (%EI/d) | 33.40 (0.31) | 32.01 (0.30) | 31.74 (0.31) | 32.83 (0.31) | 32.32 (0.31) | 0.17 |
| Proteins<sup>3</sup> (%EI/d) | 38.26 (0.39) | 36.79 (0.38) | 36.50 (0.38) | 36.22 (0.38) | 35.10 (0.38) | <.0001 |
| Carbohydrates<sup>3</sup> (%EI/d) | 97.95 (0.82) | 101.38 (0.80) | 101.91 (0.81) | 102.86 (0.81) | 105.36 (0.81) | <.0001 |
| Sugar<sup>3</sup> (%EI/d) | 16.12 (0.27) | 16.99 (0.27) | 17.10 (0.27) | 18.47 (0.27) | 20.03 (0.27) | <.0001 |
| Animal protein<sup>3</sup> (%EI/d) | 25.94 (0.40) | 24.23 (0.40) | 23.60 (0.40) | 23.44 (0.40) | 22.75 (0.40) | <.0001 |
| Plant protein<sup>3</sup> (%EI/d) | 12.90 (0.15) | 13.27 (0.15) | 13.44 (0.15) | 13.38 (0.15) | 13.00 (0.15) | 0.51 |
| SFA (g/d) | 31.47 (0.38) | 31.44 (0.38) | 31.62 (0.38) | 32.68 (0.38) | 30.83 (0.38) | 0.96 |
| Atherogenic SFA<sup>5</sup> (g/d) | 20.25 (0.25) | 20.26 (0.25) | 20.00 (0.25) | 20.78 (0.25) | 19.74 (0.25) | 0.54 |
| EPA+DHA (g/d) | 0.31 (0.02) | 0.34 (0.02) | 0.25 (0.02) | 0.25 (0.02) | 0.28 (0.02) | 0.03 |
| Linoleic FA (g/d) | 7.11 (0.13) | 6.81 (0.13) | 6.57 (0.13) | 6.89 (0.13) | 6.91 (0.13) | 0.45 |
| Alpha-linolenic FA (g/d) | 1.03 (0.03) | 1.01 (0.02) | 0.96 (0.02) | 0.92 (0.02) | 0.93 (0.02) | <.0001 |
| PUFA (g/d) | 10.08 (0.17) | 9.54 (0.17) | 9.20 (0.17) | 9.41 (0.17) | 9.75 (0.17) | 0.15 |
| Beta-carotene (µg/d) | 2737 (129) | 2563 (127) | 2890 (128) | 2715 (121) | 2445 (128) | 0.29 |
| Calcium (mg/d) | 921.50 (15.06) | 909.39 (14.86) | 913.67 (14.92) | 948.43 (14.94) | 845.54 (14.93) | 0.02 |
| Cholesterol (mg/d) | 340.33 (6.56) | 329.32 (6.47) | 315.25 (6.50) | 323.13 (6.51) | 293.95 (6.51) | <.0001 |
| Copper (mg/d) | 1.85 (0.04) | 1.76 (0.04) | 1.81 (0.04) | 1.53 (0.04) | 1.44 (0.04) | <.0001 |
| Iron (mg/d) | 10.47 (0.13) | 10.78 (0.13) | 10.58 (0.13) | 10.10 (0.13) | 10.02 (0.13) | <.0001 |
| Non-heme iron (mg/d) | 9.26 (0.11) | 9.55 (0.11) | 9.29 (0.11) | 8.85 (0.11) | 8.82 (0.11) | <.0001 |
| Zinc (mg/d) | 9.70 (0.15) | 9.76 (0.15) | 9.47 (0.15) | 9.41 (0.15) | 8.64 (0.15) | <.0001 |
| Fiber (g/d) | 20.42 (0.25) | 20.04 (0.24) | 20.12 (0.24) | 19.58 (0.24) | 18.23 (0.24) | <.0001 |
| Iodine (µg/d) | 147.58 (2.72) | 153.14 (2.68) | 148.20 (2.70) | 156.15 (2.70) | 143.66 (2.70) | 0.58 |
| Magnesium (mg/d) | 354.69 (4.26) | 355.80 (4.21) | 336.18 (4.22) | 331.81 (4.23) | 316.67 (4.23) | <.0001 |
| Manganese (mg/d) | 3.48 (0.05) | 3.22 (0.05) | 3.18 (0.05) | 2.76 (0.05) | 2.66 (0.05) | <.0001 |
| Phosphorus (mg/d) | 1,221 (11) | 1,206 (11) | 1,185 (11) | 1,179 (11) | 1,136 (11) | <.0001 |
| Phytates (µg/d) | 618.87 (9.67) | 612.97 (9.54) | 594.88 (9.58) | 579.84 (9.59) | 569.60 (9.59) | <.0001 |
| Polyols (mg/d) | 1.23 (0.06) | 1.26 (0.06) | 1.19 (0.06) | 0.97 (0.06) | 0.94 (0.06) | <.0001 |
| Potassium (mg/d) | 3,226 (35) | 3,188 (34) | 3,054 (34) | 3,028 (34) | 2,873 (34) | <.0001 |
| Retinol (µg/d) | 474.30 (33.43) | 491.88 (32.98) | 500.93 (33.11) | 435.60 (33.16) | 377.45 (33.15) | 0.02 |
| Selenium (µg/d) | 130.67 (2.04) | 123.18 (2.01) | 125.71 (2.02) | 122.92 (2.02) | 113.86 (2.02) | <.0001 |
| Sodium (mg/d) | 2,978 (43) | 3,033 (43) | 3,263 (43) | 3,273 (43) | 3,059 (43) | <.0001 |
| Vitamin B1 (mg/d) | 1.20 (0.02) | 1.20 (0.02) | 1.21 (0.02) | 1.23 (0.02) | 1.18 (0.02) | 0.87 |
| Vitamin B2 (mg/d) | 1.80 (0.03) | 1.80 (0.03) | 1.74 (0.03) | 1.83 (0.03) | 1.75 (0.03) | 0.52 |
| Vitamin B3 (mg/d) | 20.45 (0.35) | 20.56 (0.35) | 19.70 (0.35) | 19.46 (0.35) | 20.14 (0.35) | 0.13 |
| Vitamin B5 (mg/d) | 5.58 (0.07) | 5.68 (0.07) | 5.52 (0.07) | 5.60 (0.07) | 5.46 (0.07) | 0.16 |
| Vitamin B6 (mg/d) | 1.71 (0.02) | 1.71 (0.02) | 1.70 (0.02) | 1.67 (0.02) | 1.70 (0.02) | 0.41 |
| Vitamin B9 (µg/d) | 308.39 (4.24) | 306.96 (4.18) | 300.22 (4.20) | 293.86 (4.21) | 279.98 (4.20) | <.0001 |
| Vitamin B12 (µg/d) | 5.58 (0.20) | 5.64 (0.20) | 5.32 (0.20) | 4.79 (0.20) | 4.62 (0.20) | <.0001 |
| Vitamin C (mg/d) | 95.89 (2.65) | 89.08 (2.61) | 93.03 (2.62) | 85.96 (2.63) | 81.68 (2.62) | <.0001 |
| Vitamin E (mg/d) | 10.37 (0.18) | 9.71 (0.17) | 9.14 (0.17) | 9.31 (0.17) | 9.59 (0.17) | <.0001 |
| PANDiet (/100) | 59.64 (0.28) | 59.59 (0.28) | 59.08 (0.28) | 57.99 (0.28) | 56.89 (0.28) | <.0001 |
| sPNNS-GS2 (/14.25) | 1.42 (0.13) | 1.07 (0.12) | 0.55 (0.13) | 0.36 (0.13) | 0.32 (0.13) | <.0001 |

Abbreviations: EI, energy intake; DHA, docosahexaenoic acid; EPA, eicosapentaenoic acid; FA, fatty acids;
PANDiet, Diet Quality Index Based on the Probability of Adequate Nutrient Intake; PUFA, polyunsaturated
fatty acids; SFA, saturated fatty acids; sPNNS-GS2, simplified Programme National Nutrition Santé-Guidelines
Score 2
<sup>1</sup>Values are means (standard errors of the means) adjusted for energy intake using the residual method or standard adjustment for contribution of macronutrient to energy intake, PANDiet, and sPNNS-GS2.
<sup>2</sup>Values are crude means (SD)
<sup>3</sup>Values are expressed as % of total energy intake
<sup>4</sup>P-value for linear trend is estimated using linear contrast
<sup>5</sup>Atherogenic fatty acids include lauric and myristic and palmitic acids

Overall, quality of the diet was lower in Q5 compared to Q1, with a decrease in sPNNS-GS2 (−77.5%) and PANDiet (−4.6%) across quintiles (**Table 3**).

Overall, diet-related environmental pressures greatly differed depending on whether energy adjustment was applied or not (**Table 4**).

**Table 41:** Daily diet-related environmental indicators according to %UPF quintiles, (INCA 3, n=2,121) ^1^.

| Crude | Q1 | Q2 | Q3 | Q4 | Q5 | P-trend<sup>2</sup> |
| --- | --- | --- | --- | --- | --- | --- |
| Climate change (GHGe) (kg CO<sub>2</sub> eq) | 5.42 (5.15-.69) | 5.81 (5.54-6.08) | 5.96 (5.69-6.23) | 6.19 (5.92-6.45) | 6.24 (5.98-6.51) | 0.003 |
| Water use (m<sup>3</sup> world eq) | 6.90 (6.62-7.18) | 6.55 (6.27-6.83) | 6.64 (6.36-6.92) | 6.15 (5.87-6.43) | 6.44 (6.16-6.72) | 0.01 |
| Land use (pt) | 255.41 (240.34-270.48) | 272.07 (257.07-287.08) | 281.54 (266.55-296.53) | 290.87 (275.82- 305.92) | 299.58 (284.59-314.56) | 0.01 |
| Energy demand (MJ) | 58.81 (56.94-60.67) | 60.82 (58.96-62.68) | 63.34 (61.49-65.20) | 61.68 (59.82-63.55) | 63.23 (61.37-65.08) | 0.01 |
| Acidification (mol H<sup>+</sup> eq) | 0.066 (0.002) | 0.069 (0.002) | 0.071 (0.002) | 0.073 (0.002) | 0.075 (0.002) | 0.01 |
| Resource use, minerals and metals (kg Sb eq) | 9.73 (9.38-10.07) | 10.04 (9.70-10.38) | 10.00 (9.66-10.34) | 9.58 (9.24-9.92) | 9.63 (9.29-9.97) | 0.25 |
| Eutrophication, freshwater (kg P eq) | 1.02 (0.97-1.07) | 1.03 (0.98-1.08) | 1.01 (0.96-1.06) | 0.98 (0.93-1.03) | 1.02 (0.97-1.06) | 0.25 |
| Eutrophication, marine (kg N eq) | 20.96 (19.99-21.93) | 22.57 (21.61-23.54) | 23.53 (22.57-24.50) | 22.88 (21.91-23.85) | 23.61 (22.65-24.58) | 0.06 |
| Eutrophication, terrestrial (mol N eq) | 0.26 (0.25-0.28) | 0.28 (0.26-0.29) | 0.28 (0.27-0.30) | 0.30 (0.28-0.31) | 0.30 (0.29-0.31) | 0.01 |
| Photochemical ozone formation (kg NMVOC eq) | 16.19 (15.40-16.97) | 16.69 (15.91-17.47) | 16.52 (15.74-17.30) | 16.87 (16.09-17.66) | 17.00 (16.23-17.78) | 0.37 |
| Ozone depletion (kg CFC-11eq) | 0.53 (0.45-0.62) | 0.56 (0.48-0.64) | 0.51 (0.43-0.59) | 0.66 (0.58-0.74) | 0.51 (0.43-0.59) | 0.80 |
| Particulate matter (disease incidence) | 0.48 (0.46-0.50) | 0.51 (0.48-0.53) | 0.52 (0.49-0.54) | 0.53 (0.51-0.55) | 0.54 (0.52-0.56) | 0.01 |
| Ionising radiation (kBq U235 eq) | 1.27 (1.22-1.31) | 1.34 (1.30-1.38) | 1.44 (1.40-1.49) | 1.42 (1.37-1.46) | 1.47 (1.43-1.52) | <.0001 |
| EF score<sup>3</sup> | 0.66 (0.63-0.69) | 0.69 (0.67-.72) | 0.71 (0.68-0.74) | 0.71 (0.69-0.74) | 0.73 (0.70-0.75) | 0.02 |
| Adjusted for dietary energy intake | Q1 | Q2 | Q3 | Q4 | Q5 | P-trend<sup>2</sup> |
| Climate change (GHGe) (kg CO<sub>2</sub> eq) | 5.93 (5.71-6.15) | 5.84 (5.62-6.05) | 5.59 (5.37-5.80) | 5.93 (5.71-6.14) | 5.82 (5.60-6.03) | 0.19 |
| Water use (m3 world eq) | 7.34 (7.09-7.59) | 6.58 (6.34-6.82) | 6.32 (6.07-6.56) | 5.92 (5.68-6.17) | 6.07 (5.82-6.31) | <.0001 |
| Land use (pt) | 280.28 (267.28-293.28) | 273.46 (260.63-286.28) | 263.24 (250.36-276.12) | 278.17 (265.27-291.07) | 278.65 (265.76-291.54) | 0.31 |
| Energy demand (MJ) | 63.38 (62.17-64.58) | 61.07 (59.89-62.26) | 59.98 (58.79-61.17) | 59.35 (58.16-60.54) | 59.38 (58.19-60.57) | <.0001 |
| Acidification (mol H<sup>+</sup> eq) | 0.072 (0.001) | 0.070 (0.001) | 0.067 (0.001) | 0.070 (0.001) | 0.070 (0.001) | 0.11 |
| Resource use, minerals and metals (kg Sb eq) | 10.49 (10.25-10.74) | 10.08 (9.84-10.33) | 9.44 (9.19-9.68) | 9.19 (8.94-9.43) | 8.99 (8.74-9.23) | <.0001 |
| Eutrophication, freshwater (kg P eq) | 1.09 (1.05-1.13) | 1.04 (0.99-1.08) | 0.96 (0.92-1.00) | 0.94 (0.90-0.99) | 0.96 (0.91-1.00) | <.0001 |
| Eutrophication, marine (kg N eq) | 22.94 (22.19-3.69) | 22.68 (21.94-3.42) | 22.07 (21.33-2.82) | 21.87 (21.12-22.61) | 21.95 (21.20-22.69) | 0.001 |
| Eutrophication, terrestrial (mol N eq) | 0.29 (0.27-0.30) | 0.28 (0.27-0.29) | 0.27 (0.26-0.28) | 0.28 (0.27-0.29) | 0.28 (0.27-0.29) | 0.20 |
| Photochemical ozone formation (kg NMVOC eq) | 17.38 (16.69-8.07) | 16.75 (16.07-7.44) | 15.64 (14.96-16.33) | 16.26 (15.57-6.95) | 16.00 (15.31-6.69) | 0.01 |
| Ozone depletion (kg CFC-11eq) | 0.58 (0.49-0.66) | 0.56 (0.48-0.64) | 0.48 (0.40-0.56) | 0.64 (0.55-0.72) | 0.47 (0.39-0.55) | 0.39 |
| Particulate matter (disease incidence) | 0.52 (0.51-0.54) | 0.51 (0.49-0.53) | 0.49 (0.47-0.50) | 0.51 (0.49-0.53) | 0.50 (0.49-0.52) | 0.06 |
| Ionising radiation (kBq U235 eq) | 1.38 (1.35-1.41) | 1.35 (1.32-1.37) | 1.36 (1.34-1.39) | 1.36 (1.33-1.39) | 1.38 (1.35-1.41) | 0.53 |
| EF score<sup>3</sup> | 0.72 (0.70-0.74) | 0.69 (0.68-0.71) | 0.67 (0.65-0.69) | 0.68 (0.66-0.70) | 0.68 (0.66-0.70) | 0.002 |

Abbreviation: EF, ecological footprint; GHGe, greenhouse gas emissions. Units are as follows: kg CO<sub>2</sub> eq, carbon dioxide equivalent; m3 world eq, water use in cubic meters of water; land use is estimated as loss of soil organic matter content in kilograms of carbon deficit (kg C deficit) dimensionless and expressed as Points (Pt); MJ, megajoule; mol H<sup>+</sup> eq, equivalent of moles hydron; kg Sb eq, equivalent of kilograms of antimony; kg P eq, equivalent of kilograms of phosphorus, kg N eq, equivalent of kilograms of nitrogen; mol N eq, equivalent of moles of nitrogen; kg NMVOC eq, equivalent of kilograms of non-methane volatile organic compounds; kg CFC-11eq, equivalent of kilograms of trichlorofluoromethane (Freon-11); Emission of particulate matter in change in mortality due to particulate matter emissions; kg U235 eq, equivalent of kilobecquerels of Uranium 235
<sup>1</sup>Values are means (95%CI) (crude or energy-adjusted)
<sup>2</sup>P-value for linear trend is estimated using contrast
<sup>3</sup>For the EF score, the higher it is, the more impactful it is

In the unadjusted model, most indicators were higher among participants with higher %UPF, including GHGe (Q5 vs. Q1: +15%), land use (+17%), fossils resource use (8%), marine and terrestrial eutrophication (+13% and 15%), particulate matter (+13%), ionizing radiation (+16%) and the overall endpoint ecological footprint (EF) score (+11%). On the contrary, water use was inversely associated with %UPF with a lower mean value found in Q5 compared to Q1 (−7%).

When adjustment for energy intake was applied, some associations were no longer significant, including GHGe while the negative association with regard to water use was slightly strengthened (Q5 vs. Q1 %UPF=-17%). In addition, associations regarding resource use, freshwater and marine eutrophication, and ozone formation were reversed as well as the association concerning the global EF score (−6%).

GHGe, land use, energy demand and water use, according to quintiles of %UPF are presented by food supply chain stages in **Figure 1** and **Supplemental Table 2** (energy-adjusted models). Overall, whatever the quintile, agricultural production of raw material was the main driver for GHGe, land use and water use, while packaging and processing stages were also important contributors to the energy demand pressure. The substantial differences between %UPF quintiles (Q5 vs. Q1) were higher impacts of the processing stage on water use (+53%), GHGe (+42%) and energy demand (+37%).

**Figure 1:**
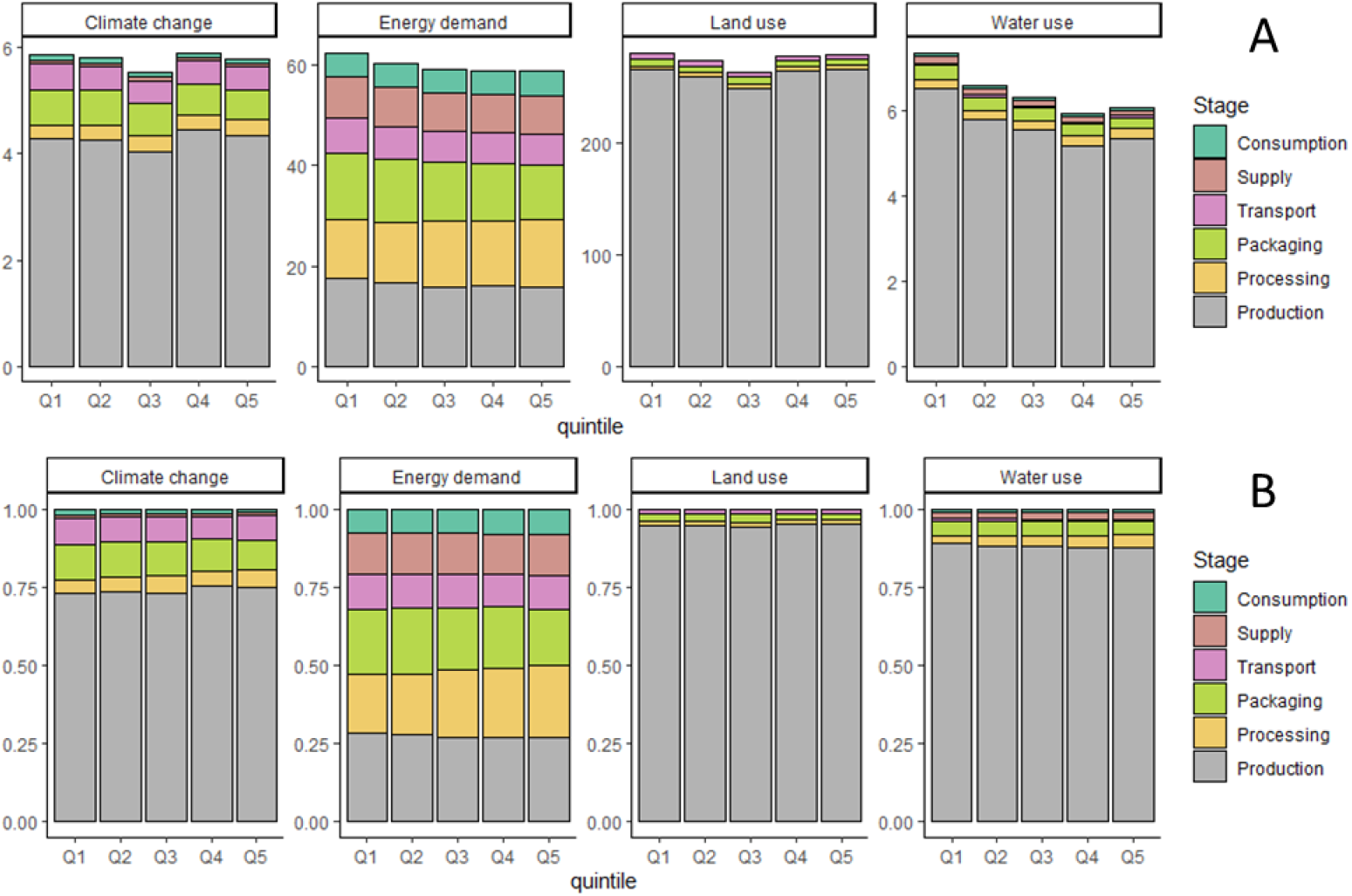
Farm and post-farm stages contribution to daily diet-related environmental indicators according to %UPF quintiles (INCA 3, n=2,121), adjusted for energy intake ^1,2^. Panel A corresponds to the pressures, in absolute values, of each stage. Panel B corresponds to the pressures, in relative value (%), of each stage ^1^Climate change (Greenhouse gas emissions), energy demand, land use and water use are expressed in kg CO2eq, m^3^ world eq, pt, MJ, respectively ^2^Energy-adjusted values

Concerning results by NOVA class (**Figure 2** and **Supplemental Table 3**), NOVA1 (unprocessed) food consumption greatly contributed to each environmental indicator in all quintiles. However, the contribution of UPF consumption to environmental pressures greatly differed according to quintile and indicators. In Q1, GHGe, land use, energy demand and water use related to consumption of UPF contributed to 10% to 13% while in Q5, contributions of NOVA4 consumption were 35%, 39%, 28% and 42% to total GHGe, water use, land use, and energy demand, respectively.

**Figure 2:**
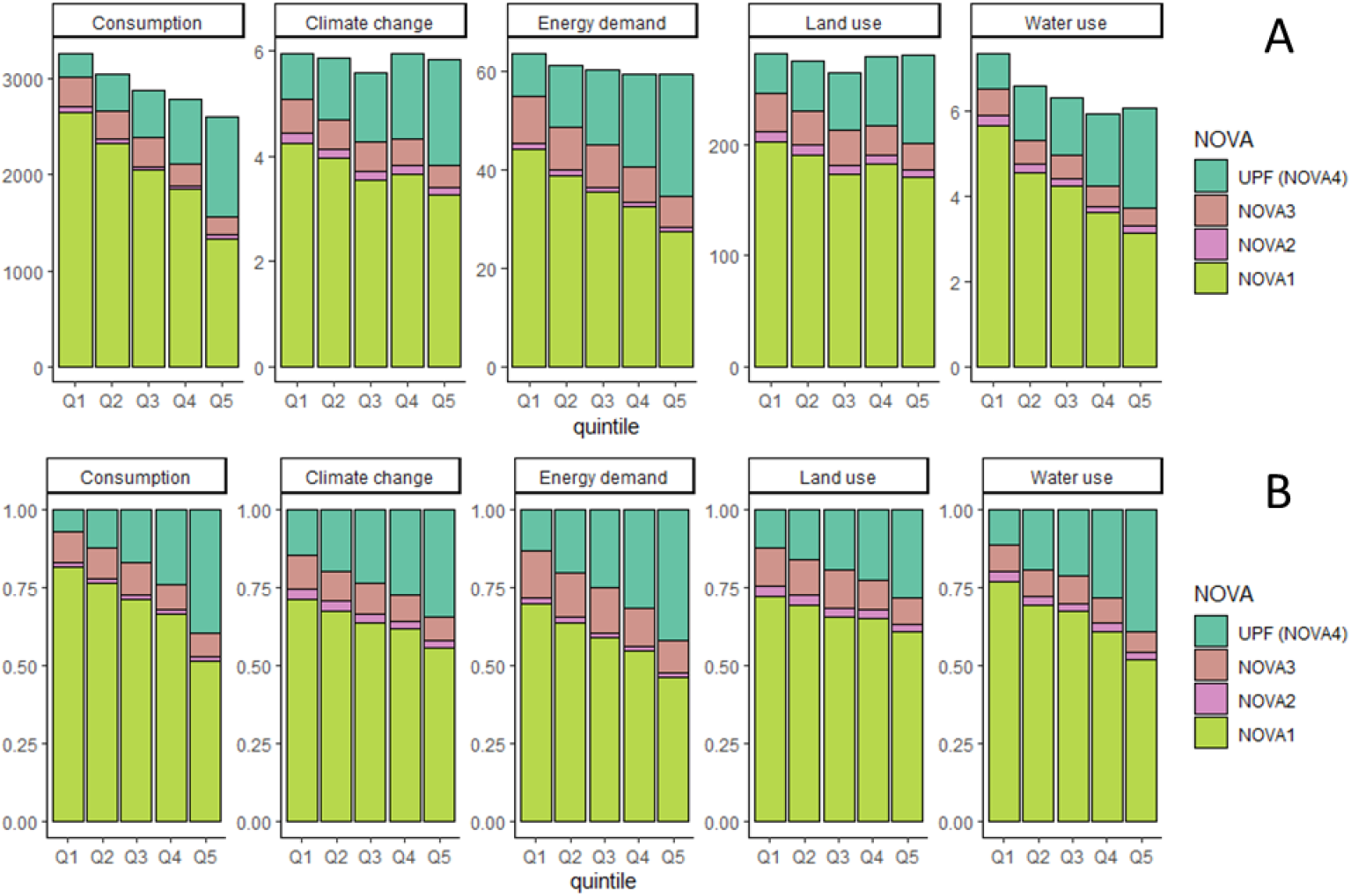
Contribution of NOVA class consumption to daily diet-related environmental indicators according to %UPF quintiles (INCA 3, n=2,121), adjusted for energy. Panel A corresponds to the pressures, in absolute value, related to %NOVA class in the diet. Consumption, climate change (Greenhouse gas emissions), energy demand, land use and water use are expressed in g/d, kg CO2eq, m^3^ world eq, pt, MJ, respectively. Panel B corresponds to same data but in relative value (%).

In sensitivity analyses, further adjustment for %NOVA1 in addition to energy intake (**Supplemental Table 4**), yielded strengthened associations, with notably increases in GHGe (Q5 vs. Q1=+32%) and land use (Q5 vs. Q1=+29%).

With the substitution model from NOVA1 (unprocessed) to UPF consumption (modeled as a continuous variable) produced similar trends to the models adjusted for energy intake (**Figure 3**); with a decrease in water use and an increase in GHGe and land use from production and high increase in energy demand from processing.

**Figure 3:**
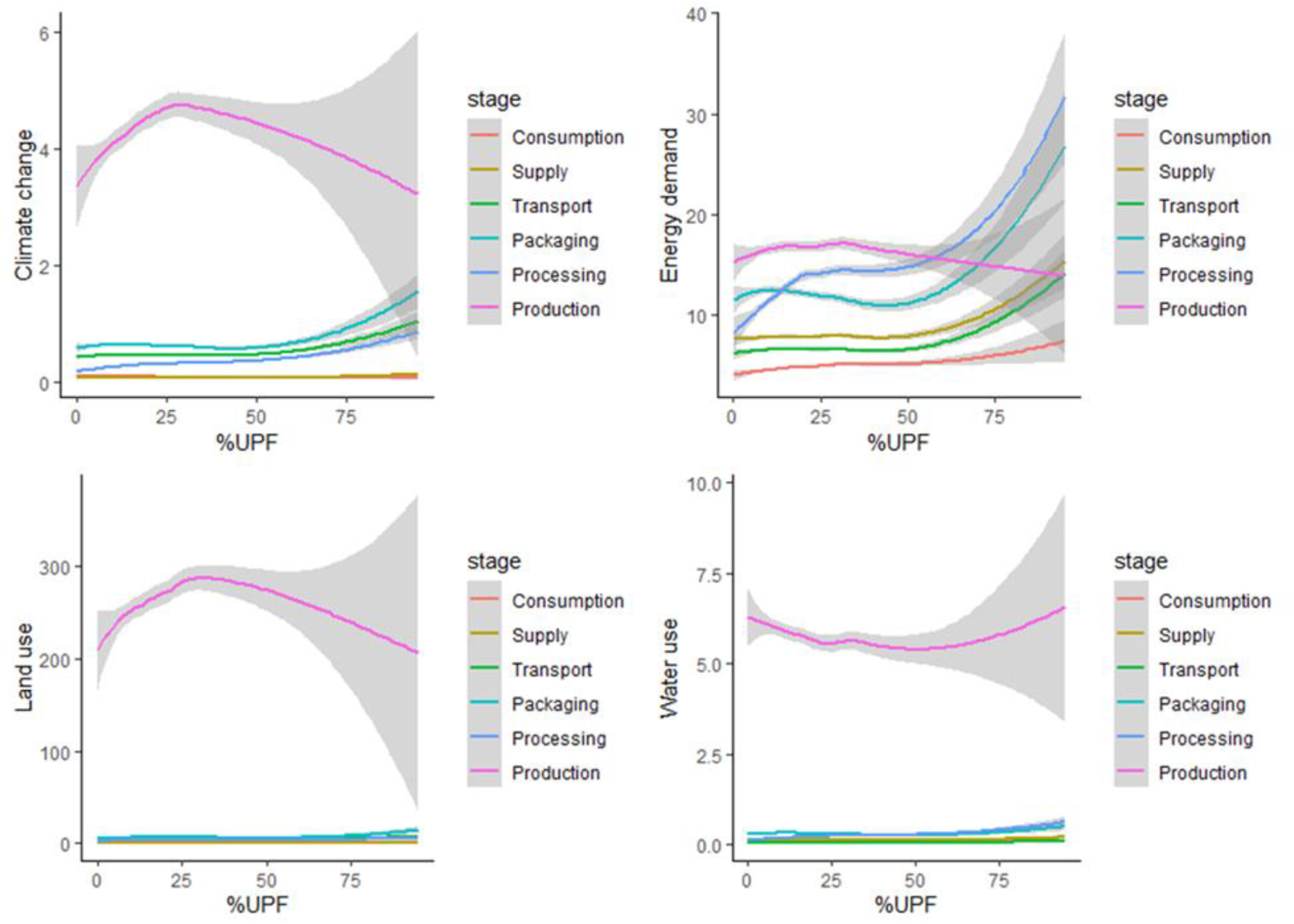
Daily diet-related environmental indicators for each step for substitution model of UPF (INCA 3, n=2,121)^1,2^. ^1^ Climate change (Greenhouse gas emissions), energy demand, land use and water use are initially expressed in g/d, kg CO2eq, m^3^ world eq, pt, MJ, respectively ^2^ Model is substitution of NOVA1 (unprocessed) by UPF adjusted for energy intake

Finally, results regarding % UPF as calorie are presented in **Supplemental Table 5**. Overall, the results were similar but trend across quartiles was less evident. Notably, the association between GHGe and level in UPF in the diet remained in the energy-adjusted model (+11%).

## Discussion

In the present study conducted in a representative French population-based survey, we observed that most of the studied environmental pressure indicators were higher among participants with higher %UPF in their diet, in particular GHGe, energy demand and land use. A large part of the higher pressures observed was explained by higher energy intake. Most environmental pressures occurred at the stage of agricultural production, apart from energy demand for which the processing and packaging stages was also an important contributor. NOVA1 food consumption highly contributed to land use and GHGe in all quintiles. NOVA4 food consumption greatly contributed to energy demand and to a lesser extent to water use with a strong gradient across quintiles.

To our knowledge, only one study previously investigated the environmental pressure of diet according to the degree of food processing evaluated through the NOVA classification (20). Comparison of our results with those of this study is not straightforward, since it has been conducted in a Brazilian population, which may have exhibited some dissimilarities as regards dietary habits and environmental pressures associated to food production and processing. As found herein, this study reported difference in energy intake and GHGe according to level of UPF in the diet. Also, in our study, the positive relationship between GHGe and %UPF did not remain significant after adjustment for energy. In the study by Garzillo *et al* (20), the singular role of energy intake was not available as the authors also adjusted for sociodemographic factors, which may be considered questionable inasmuch as this is not supposed to confuse the relation when focusing on the link between dietary patterns and environmental pressures. Moreover, when %UPF was expressed in kcal, as performed in the study of Garzillo *et al*., the associations between GHGe and quintiles of %UPF remained statistically significant even in the energy-adjusted model. Besides, our findings related to water footprint were not similar to those observed in the Brazilian study, since they documented a higher food-related water use in participants with higher levels of UPF in their diet, which did not remain after energy intake adjustment. In contrast, in our study, a higher water footprint from diet of participants was observed among low %UPF participants, the association was even strengthened after adjustment for energy intake. This latter result was attributable primarily to the fact that participants who consumed lower amounts of UPF and, thus, in proportion, more NOVA1 food, tended to have higher fruit and vegetable intakes and non-alcoholic beverages. This is also clearly observed in our substitution model modelled with %UPF as a continuous variable. Thus, our finding is consistent with the literature documenting a higher water footprint in plant-rich diets (37). Similarly, at constant energy intake, the fact that the association between %UPF in the diet and GHGe and land use disappeared, is related to the somewhat higher red meat, dairy products, fruit, vegetables, and fish consumption (all classified as NOVA1) among low consumers of UPF compared to high consumers, as previously reported (5). Of note meat, in particular ruminant meat, is the strongest contributor to GHGe (17,38). These findings were consistent with the strong increase in environmental pressure with %UPF when adjustment for NOVA1 was performed.

The present work is the first to explore the pressures associated with the production stage and all post-farm stages including processing, packaging, transport and supply, according to the degree of processing in the diet. Our results showed an important role of post-farm stages in energy demand whatever the %UPF in the diet. The agricultural production stage is the main contributor to diet-related GHGe, land and water use while with regard to energy demand, this is less evident since post-farm stages play also an important role in the total pressure.

Our findings indicate that environmental impacts of UPF consumption could be linked to at least two main factors. First, the higher energy intake of high consumers of UPF is a major determinant of their diet-related impacts (39,40). Second, the higher number of post-farming stages for UPF production can lead to an increase in energy demand and various environmental pressures. Indeed, UPF are, by definition, related to more industrial processing, more packaging (for instance, they represent 2/3 and more than 70% of packaged foods in France and in the US, respectively), and longer transport which may substantially contribute to environmental impact of food (41). Consistently with the scientific literature (16,17), even if the production stage is the most impacting stage for the environmental resources depletion, it appeared that the food processing consumes a large amount of fossil resources. On the contrary, it has been suggested that UPF (on a 100kcal basis) are less GHG emitting and environmentally harmful (water and land use) than some minimally/unprocessed foods, in particular if they contain small amount of animal ingredients (14,17,42). This last aspect is consistent with 1) our results when dietary energy intake was accounted for, as most the pressures decreased across the quintiles, and 2) with a recent British study analyzing foods properties that documented that processed foods have lower nutritional quality, but also lower GHGe and were less expensive than minimally processed foods, regardless of their total fat, salt and/or sugar content (42).

Studies that have explored the environmental pressures of UPF in details are very limited and none of them considered the details of the post-agricultural stages, even though this element is essential to accurately assess environmental impacts of such foods. In addition to potential human health benefits (4,7–13), the reduction of high UPF consumption, associated with a greater overall consumption, could be a driver in the transition towards a more sustainable food system by contributing to the reduction of GHGe, energy demand, land use, soil and water degradation, and pollution. In addition, it has been documented that production of UPF is also associated with the use of fertilizers and pesticides, deforestation and biodiversity loss as well as packaging (2,19,43,44). With respect to our findings as regards water use, the benefit is less convincing since, as previously emphasized (37), healthy diets rich in fruits, vegetables and nuts are water consuming. Thus, some discrepancies exist in alignment of foods as regards environmental sustainability and health impacts. For instance, it is now well known that sugar, salt and food staples can have lower environmental impacts per calorie than fruits, vegetables, and animal-based foods (45,46). The main lever to achieve food sustainability remains the reduction of red meat and processed meat intake, as this would benefit for both human health and the environment (14).

Our study exhibits some limitations and strengths. First, the study was based on diets of French adults in a relatively small – though representative – sample, limiting the diversity of dietary patterns such as vegetarian diets. For example, participants in INCA 3 had an average age of 47 years, whereas consumers of UPF have been shown to be young (47). Second, limitations were somewhat inherent to the matching between Agribalyse 3.01 and INCA 3, as food databases were independently developed, and environmental indicators were not available for certain foods (for instance, culinary aids) or not detailed (for instance, type of mushrooms). Third, even though the Agribalyse database is very rich and accurate, some elements have been prioritized. For example, regarding packaging, only B2C (business-to-consumer) packaging was considered and not B2B (business-to-business). Regarding processing, the focus was driven by stages related to mass and yield changes and some other lacking have been underlined (35). As regards transport, average values along the value chain were considered but transport from the supply point to the household was not considered. Finally, the data were based on LCA according to the standardized guidelines and methodologies but did not consider the type of farming system (organic or conventional), limiting the consideration of the variety of practices along the food chain. In addition, some indicators such as biodiversity loss were not available. As to the strengths of our study, dietary data was collected in a nationally representative sample of the adult population of France in 2015-2016. Furthermore, the consumption and pressures data were collected using standardized methodologies. In addition, environmental data were validated by several expert entities (35). Finally, the detail of pressures by stage of the value chain allowed us to consider for the first time the contribution of UPF while taking into the different stages.

## Conclusion

This study is the first to explore the contribution of UPF consumption to different environmental pressures, while detailing the different stages of the food chain. As high consumers of ultra-processed foods generally have higher energy intake, overall their diets were associated with a higher footprint. In addition, the consumption of UPF had a substantial role on some indicators, particularly on energy demand through the processing stage. Such investigations could be considered in the development of sustainable dietary guidelines in light of the previously documented links between UPF and human health.

## Data Availability

Data availability: Data described in the manuscript are available at: https://www.data.gouv.fr/fr/datasets/donnees-de-consommations-et-habitudes-alimentaires-de-letude-inca-3/.

## The authors’ contributions

EKG conducted the research, implemented the databases, conducted the analyses and wrote the manuscript.

All authors critically helped in the interpretation of results, revised the manuscript and provided relevant intellectual input. They all read and approved the final manuscript.

EKG had primary responsibility for the final content, she is the guarantor.

## Acknowledgment

The authors are indebted to Vincent Colomb and Mélissa Cornélius (ADEME) for their support in the use of the Agribalyse database ®.

