## Supplemental materials for "Environmental impacts associated with UPF consumption: which food chain stages matter the most? *Findings from a representative sample of French adults*"

**Online supplementary material**
*Kesse-Guyot et al.*

**Supplemental Material 1: NOVA classification**

The methodology for the classification of foods according to NOVA presented in this supplemental has been described in a previous publication (1). All food and beverage items of the INCA 3 composition table (n>2,800) were categorized into one of the four NOVA groups, a food classification system based on the extent and purpose of industrial food processing (2–4). The "ultra-processed foods" (UPF) group of the NOVA classification is the primarily focus of this study. Products in this group undergo industrial processes that include for instance hydrogenation, hydrolysis, extruding, molding, reshaping, and pre-processing by frying. Flavoring agents, colors, emulsifiers, humectants, non-sugar sweeteners and other cosmetic additives are often added to these products to imitate sensorial properties of unprocessed or minimally processed foods and their culinary preparations. The UPF group is defined by opposition to the other NOVA groups: "unprocessed or minimally processed foods" (fresh, dried, grounded, chilled, frozen, pasteurized or fermented staple foods such as fruits, vegetables, pulses, rice, pasta, eggs, meat, fish or milk), "processed culinary ingredients" (salt, vegetable oils, butter, sugar and other substances extracted from foods and used in kitchens to transform unprocessed or minimally processed foods into culinary preparations) and "processed foods" (canned vegetables with added salt, sugar-coated dry fruits, meat products only preserved by salting, cheeses and freshly made unpackaged breads, and other products manufactured with the addition of salt, sugar or other substances of the "processed culinary ingredients" group). As previously described (5), home-made and artisanal food preparations were identified and decomposed using standardized recipes, and the NOVA classification was applied to their ingredients. Examples of such products as well as examples of distinctions between ultra-processed products and products from other NOVA categories are provided below:

Examples of typical ultra-processed foods according to the NOVA classification:

*Poultry and fish nuggets and sticks and other reconstituted meat products transformed with addition of preservatives other than salt (e.g. nitrites); instant noodles and dehydrated soups; carbonated diet and regular sodas; chocolate with emulsifiers, chewing gums and candies with dyes (confectionery); margarine; instant desserts; most breakfast 'cereals', 'energy' bars; 'energy' drinks; flavored milk drinks; sweet desserts made from fruit with added sugars, artificial flavors and texturizing agents; cooked seasoned vegetables with ready-made sauces; vegetable patties (meat substitutes) containing food additives; 'health' and 'slimming' products such as powdered or 'fortified' meal and dish substitutes.*

For instance, salted-only red or white meats are considered as "processed foods" whereas smoked or cured meats with added nitrites and conservatives, such as sausages and ham are classified as "ultra-processed foods".

Similarly, canned salted vegetables are considered as "processed foods" whereas industrial cooked or fried seasoned vegetables, marinated in industrial sauces with added flavorings are considered as "ultra-processed foods".

Flavored breakfast cereals with added emulsifiers, texturizing agents and/or colorants were included in the ultra-processed food group. Homemade granola, oatmeal, rye and barley flakes without additives were not considered as ultra-processed.

Regarding soups, canned liquid soups with added salts, herbs and spices are considered as "processed foods" while instant dry soup mixes are considered as "ultra-processed foods".

Example of list of ingredients for an industrial chicken and leek flavor soup considered as "ultra-processed" according to the NOVA classification: *"Dried Glucose Syrup, Potato Starch, Flavorings, Salt, Leek Powder (3.6%), Dried Leek (3.5%), Onion Powder, Dried Carrot, Palm Oil, Dried Chicken (0.7%), Garlic Powder, Dried Parsley, Colour [Curcumin (contains MILK)], Ground Black Pepper, MILK Protein, Stabilisers (Dipotassium Phosphate, Trisodium Citrate)".*

**Examples of food products considered as ultra-processed according to the NOVA classification**

| **Ultra-processed food group** | **Examples of foods** |
|---|---|

| | |
|---|---|
| Beverages | Sugary drinks (e.g. regular sodas, sugary fruit-based and flavored beverages, industrial chocolate powder beverages, energy drinks, flavored waters); artificially sweetened beverages (e.g. diet sodas, artificially sweetened ice teas) |
| Dairy products | Flavored or artificially sweetened yoghurts; products such as dairy desserts, cream cheese, milkshakes, dairy beverages, flavored milk with one or more texturizer, emulsifier, colorant or other cosmetic additives |
| Fats and sauces | Sauces and dressings (salad dressing, mayonnaise, ketchup, béchamel, and other dressings) containing emulsifiers, texturizers, flavor enhancers or other additives |
| Fruits and vegetables | Instant powder soups; reconstituted vegetarian/soy steaks with additives; flavored and artificially sweetened fruit compotes; vegan nuggets |
| Meat, fish, and eggs | Processed meat with added nitrites; chicken nuggets; fish fingers; industrial 'cordon bleu' chicken with wheat dextrose, emulsifiers, preservatives; surimi-crab sticks |
| Starchy foods and cereals | Flavored breakfast cereals with added emulsifiers, texturizing agents and/or colorants; industrial pre-baked breads and buns with added dextrose, preservatives or emulsifiers. |
| Sugary products | Industrially packed cookies, cakes, chocolate/wafer bars, and candies manufactured with glucose syrup, modified starch, hydrogenated oils, colors, flavors, emulsifiers |
| Salty snacks | Chips, crisps and crackers made with other ingredients than potatoes, oil and salt such as maltodextrin, flavors, dyes, emulsifiers, flavor enhancers |

**Supplemental Material 2: Dietary indexes computation**

The nutritional quality of the diets was assessed using two dietary indexes.

The nutrient-based PANDiet (Diet Quality Index Based on the Probability of Adequate Nutrient Intake) contains two subscales reflecting adequacy and moderation (6,7). For each nutrient, the 'probability of adequacy', i.e. intake above minimum values (adequacy score) or below maximum values (moderation score) is calculated on the basis of nutrient reference values. The final score is the average of the two sub-scores. The adequacy sub-score is the average of the probabilities of adequacy for 27 nutrients and the moderation sub-score includes 6 nutrients and 12 penalty values referring to the probabilities of exceeding upper limits of intakes. The PANDiet ranges from 0 to 100 points, with a higher score reflecting better adherence to French nutritional recommendations and adequate nutrient intake. The calculation to estimate the adequacy of the usual intake for a given nutrient is as follows:

$$Prob(\frac{y-r}{SDr})$$

Where Prob: is the *probnorm* function of SAS®, y: daily mean intake, r: the reference value, SDr: the interindividual variability.

The sPNNS-GS2 is a validated score, ranging from -∞ to 14.25, reflecting adherence to the 2017 French food-based dietary guidelines proposed by the High Council of Public Health (8,9). It is composed of 12 weighted components for moderation or adequation. The sPNNS-GS2 consists of 6 adequacy components and 6 moderation components, based on epidemiological evidence. The components are weighted according to the level of evidence for the associations with health, and finally, a penalty on energy intake is applied for over consumers. The sPNNS-GS2 includes components related to fruit and vegetables, legumes, whole grain, nuts, fish, red meat, processed meat, sweet products, sweet beverages, added lipids, alcohol, dairy products, and salt.

**Supplemental Material 3: Agribalyse database**

The diet-related environmental pressures were estimated by indicators resulting in matching consumption with the French database Agribalyse® 3.0.1 developed by the French Agency for the Environment and Energy Management (ADEME) and allowing a matching with the CIQUAL French food composition table (10).

Environmental indicator estimations are based on the method of Life Cycle Assessment (LCA) whose scope is "from field to plate". The perimeter of the indicators covers each process of the value chain: agricultural production, transport, processing, packaging, distribution and retailing, preparation at the consumer's and disposal of packaging and these processes have been split into two phases 1) production and 2) post-farm. Of note, losses and wastes (other than the non-edible parts) at home as well as transport from the retail to the household have not been considered.

Overall, the method is based on the international LCA standards: ISO 14040 (11) and ISO 14044(12), LEAP guidelines (13) and product environmental footprint (PEF) (14) and the finalized indicators are provided per kg of product and are detailed per process.

For the agricultural phase of plant products, all upstream processes (notably input production) except storage or drying are included except for ingredients used in the case of processed food. In the case of animal products, all operations including the phases of production, transport and storage of feed, fattening of animals, milking, construction and maintenance of buildings and machinery have been considered. The scope chosen is consistent with those defined in GESTIM (15) and ecoinvent® (16).

The LCI (life cycle inventory) data of AGRIBALYSE v3.0.1 covered the period 2005-2009, except for perennial crops (2000-2010). The variety of production systems was considered by applying coefficients based on the share of systems in national production. The allocation rules are varied and are based on international recommendations as described by the ISO 14040/14044 standards (11,12). In particular, allocations have been developed in order to distribute organic nitrogen fertilizers and mineral fertilizers (P and K) between crop sequences. Biophysical allocations were used for animal production (milk versus meat). The biophysical models used for animal production and allocations by type of productions are presented in the full report (17) according to the reference AFNOR-BPX 30-323 (18)(AFNOR, 2011) and in compliance with the ISO 14044 standard (12) according to 3 rules in descending order: 1) avoid allocation, 2): biophysical allocation and 3) economic allocation.

A characterization method recommended by the European Commission (Environmental Footprint 3.0) translates the input and output flows of the inventory into impacts. For the background data (inputs in construction, raw materials, etc.) the ecoinvent® database is used to assess the indirect emissions (off-field emissions). The full methodology and methodological choices have been already described (17).

The transition from commodities to food as consumed introduced coefficients related to the edible part and economic allocations between co-products. The recipes were disaggregated into ingredients, for feasibility reasons, a threshold of 95% of the ingredients covered was used. Similarly, for the origin of the ingredients, a threshold of 70% coverage was used followed by a standardization step. The whole methodology and methodological adoptions have been described elsewhere (19) and post-farm estimations are aligned with the PEF guidelines (14)

A total of 14 midpoint indicators are available: climate change, ozone depletion, particulate matters, ionizing radiation (effect on human health), ecotoxicity, photochemical ozone formation (effect on human health), acidification, terrestrial eutrophication, freshwater eutrophication, marine eutrophication, land use, water use, minerals and metals resource use, and fossils resource use.

In addition, the EF 3 (environmental footprint) single score including 2 further indicators related to human toxicity is provided. The normalization and weighting factors considered in the calculation of the EF 3 score have been extensively described (14).

Concerning the indicators for food as consumed, and according to the guidelines of the PEF method (14), a quality indicator (DQR) is provided and the AGRIBALYSE 3.0 database has been reviewed and criticized by RIVM and GreenDelta as well as by French agricultural and agri-food technical institutes "Peter Koch Consulting".

**Supplemental Table 1: Consumption of food groups (g/d) according to %UPF quintiles (in weight), (INCA 3, N=2,121)[1]**

| | Q1 | Q2 | Q3 | Q4 | Q5 | *ΔQ5 vs. Q1*[2] | P for trend[3] |
|---|---|---|---|---|---|---|---|
| Alcoholic beverages | 77.3 (112.0) | 129.8 (228.4) | 143.9 (219.9) | 124.9 (234.6) | 165.8 (377.7) | *114* | <.0001 |
| Sweetened beverages | 26.7 (101.0) | 41.1 (93.9) | 47.0 (89.9) | 119.4 (164.2) | 318.4 (449.9) | *1093* | <.0001 |
| Non-sweetened beverages | 1747.1 (737.2) | 1580.6 (686.9) | 1422.4 (618.6) | 1254.7 (633.4) | 941.1 (535.2) | *-46* | <.0001 |
| Sweet and fat foods | 81.4 (70.2) | 111.1 (82.6) | 115.3 (91.5) | 127.9 (102.0) | 129.6 (93.1) | *59* | <.0001 |
| Condiments | 22.9 (20.5) | 25.3 (26.9) | 28.7 (29.2) | 30.9 (42.0) | 26.4 (31.5) | *15* | 0.01 |
| Whole grains | 7.2 (18.5) | 7.6 (25.2) | 7.6 (24.2) | 5.8 (17.8) | 4.4 (18.3) | *-39* | 0.02 |
| Refined cereals | 186.5 (127.3) | 182.1 (114.3) | 211.2 (142.0) | 169.5 (104.6) | 173.8 (112.0) | *-7* | 0.04 |
| Fruits | 180.5 (149.8) | 161.0 (141.9) | 152.0 (133.3) | 119.9 (126.9) | 105.4 (122.4) | *-42* | <.0001 |
| Fruit juice | 44.3 (78.4) | 57.5 (95.5) | 68.8 (98.6) | 67.2 (94.1) | 81.3 (124.4) | *84* | <.0001 |
| Vegetables | 181.0 (127.6) | 157.6 (125.0) | 142.9 (100.5) | 124.5 (103.9) | 99.2 (93.1) | *-45* | <.0001 |
| Legumes | 4.6 (15.7) | 6.9 (24.3) | 6.3 (17.8) | 11.4 (28.6) | 9.8 (34.6) | *112* | <.0001 |
| Soup | 30.5 (81.4) | 60.3 (107.9) | 115.1 (159.2) | 152.1 (234.4) | 143.2 (225.2) | *369* | <.0001 |
| Potatoes | 41.5 (62.8) | 41.2 (57.5) | 52.0 (89.1) | 45.5 (61.8) | 55.8 (89.8) | *35* | 0.004 |
| Nuts | 3.7 (9.4) | 3.2 (8.8) | 3.5 (9.0) | 3.1 (8.6) | 2.9 (9.0) | *-22* | 0.21 |
| Animal fat | 9.6 (11.6) | 8.9 (13.3) | 10.5 (15.0) | 10.2 (16.3) | 8.8 (12.3) | *-9* | 0.82 |
| Vegetable fat | 9.6 (10.8) | 8.3 (9.3) | 7.2 (8.4) | 7.7 (9.6) | 8.8 (11.3) | *-8* | 0.16 |
| Prepared dishes | 101.1 (98.6) | 110.2 (110.8) | 110.5 (104.2) | 126.2 (129.7) | 132.8 (124.0) | *31* | <.0001 |
| Dairy products | 173.8 (142.6) | 186.8 (217.4) | 194.0 (155.0) | 210.9 (171.3) | 170.3 (176.6) | *-2* | 0.52 |
| Eggs | 17.5 (27.4) | 14.8 (30.3) | 11.0 (22.3) | 13.9 (25.6) | 12.0 (24.3) | *-31* | 0.003 |
| Fish | 34.7 (44.0) | 29.8 (36.6) | 25.7 (38.4) | 24.8 (40.0) | 26.2 (40.4) | *-25* | 0.0003 |
| Red meat | 50.3 (50.3) | 52.6 (56.8) | 53.6 (57.6) | 52.3 (59.3) | 50.8 (62.9) | *1* | 0.93 |
| Processed meat | 23.3 (24.7) | 29.0 (33.5) | 35.7 (41.2) | 33.5 (46.9) | 35.2 (45.7) | *51* | <.0001 |
| Poultry | 27.1 (33.8) | 26.3 (38.7) | 30.1 (37.4) | 23.2 (33.3) | 28.7 (39.3) | *6* | 0.99 |

[1]Values are unadjusted means (standard deviation)

[2]Relative difference

[3]P-value for linear trend is estimated using linear contrast

**Supplemental Table 2: Environmental indicators according to %UPF quintiles (in weight) according to farm and post-farm stages, (INCA 3, n=2,121)[1]**

| Variable | Variable | Q1 | Q2 | Q3 | Q4 | Q5 | P for trend[2] |
|---|---|---|---|---|---|---|---|
| Climate change | Production | 4.28 (4.07-4.48) | 4.27 (4.06-4.47) | 4.05 (3.85-4.26) | 4.45 (4.24-4.66) | 4.34 (4.13-4.54) | 0.37 |
| (GHGe) | Processing | 0.26 (0.25-0.27) | 0.27 (0.26-0.27) | 0.29 (0.28-0.30) | 0.28 (0.27-0.29) | 0.32 (0.31-0.32) | <.0001 |
| (kg CO2 eq) | Packaging | 0.67 (0.65-0.69) | 0.65 (0.63-0.67) | 0.60 (0.58-0.62) | 0.59 (0.56-0.61) | 0.55 (0.53-0.57) | <.0001 |
| | Transport | 0.48 (0.47-0.49) | 0.46 (0.45-0.47) | 0.44 (0.43-0.45) | 0.43 (0.42-0.44) | 0.44 (0.43-0.45) | <.0001 |
| | Supply | 0.07 (0.07-0.07) | 0.06 (0.06-0.06) | 0.06 (0.06-0.06) | 0.06 (0.06-0.06) | 0.06 (0.06-0.06) | <.0001 |
| | Consumption | 0.11 (0.11-0.11) | 0.09 (0.09-0.10) | 0.09 (0.09-0.09) | 0.08 (0.07-0.08) | 0.07 (0.07-0.08) | <.0001 |
| Water use | Production | 6.52 (6.28-6.77) | 5.80 (5.56-6.04) | 5.55 (5.31-5.79) | 5.17 (4.93-5.41) | 5.33 (5.09-5.57) | <.0001 |
| ($m^3$ world eq) | Processing | 0.19 (0.18-0.20) | 0.19 (0.18-0.20) | 0.21 (0.20-0.22) | 0.22 (0.21-0.23) | 0.24 (0.23-0.25) | <.0001 |
| | Packaging | 0.34 (0.33-0.35) | 0.32 (0.31-0.33) | 0.30 (0.29-0.31) | 0.28 (0.27-0.29) | 0.26 (0.25-0.27) | <.0001 |
| | Transport | 0.05 (0.05-0.05) | 0.05 (0.04-0.05) | 0.04 (0.04-0.04) | 0.04 (0.04-0.04) | 0.04 (0.04-0.04) | <.0001 |
| | Supply | 0.15 (0.15-0.15) | 0.14 (0.14-0.15) | 0.14 (0.13-0.14) | 0.13 (0.13-0.14) | 0.13 (0.12-0.13) | <.0001 |
| | Consumption | 0.08 (0.08-0.08) | 0.07 (0.07-0.08) | 0.07 (0.07-0.07) | 0.07 (0.06-0.07) | 0.07 (0.06-0.07) | <.0001 |
| Land use | Production | 265.05 (252.04-278.07) | 258.75 (245.91-271.59) | 248.05 (235.16-260.94) | 264.47 (251.56-277.39) | 265.41 (252.50-278.31) | 0.76 |
| (pt) | Processing | 3.81 (3.58-4.05) | 3.68 (3.45-3.91) | 3.99 (3.76-4.23) | 3.76 (3.52-3.99) | 3.73 (3.50-3.97) | 0.83 |
| | Packaging | 6.61 (6.37-6.85) | 6.55 (6.31-6.79) | 6.88 (6.64-7.12) | 5.81 (5.57-6.05) | 5.51 (5.27-5.75) | <.0001 |
| | Transport | 4.18 (4.09-4.27) | 3.99 (3.90-4.07) | 3.83 (3.74-3.91) | 3.71 (3.62-3.79) | 3.63 (3.54-3.71) | <.0001 |
| | Supply | 0.21 (0.20-0.21) | 0.20 (0.19-0.20) | 0.19 (0.19-0.20) | 0.19 (0.18-0.19) | 0.19 (0.19-0.19) | <.0001 |
| | Consumption | 0.15 (0.14-0.15) | 0.14 (0.14-0.14) | 0.14 (0.13-0.14) | 0.13 (0.13-0.13) | 0.14 (0.13-0.14) | <.0001 |
| Energy demand | Production | 17.58 (17.05-18.11) | 16.77 (16.25-17.30) | 15.88 (15.35-16.41) | 15.93 (15.40-16.46) | 15.88 (15.35-16.41) | <.0001 |
| (MJ) | Processing | 11.75 (11.33-12.17) | 11.84 (11.42-12.25) | 13.03 (12.61-13.45) | 12.92 (12.50-13.34) | 13.42 (13.00-13.83) | <.0001 |
| | Packaging | 13.12 (12.72-13.53) | 12.60 (12.20-13.00) | 11.68 (11.28-12.08) | 11.47 (11.07-11.87) | 10.70 (10.30-11.11) | <.0001 |
| | Transport | 6.96 (6.82-7.11) | 6.64 (6.50-6.78) | 6.39 (6.25-6.53) | 6.24 (6.10-6.39) | 6.27 (6.13-6.42) | <.0001 |
| | Supply | 8.22 (8.03-8.40) | 7.82 (7.64-8.01) | 7.59 (7.40-7.77) | 7.53 (7.34-7.71) | 7.63 (7.44-7.81) | <.0001 |
| | Consumption | 4.73 (4.58-4.88) | 4.71 (4.56-4.86) | 4.66 (4.51-4.80) | 4.67 (4.52-4.81) | 4.91 (4.76-5.05) | 0.21 |

Abbreviations: GHGe, greenhouse gas emissions

[1]Values are energy-adjusted (95%CI)

[2]P-value for linear trend is estimated using linear contrast

**Supplemental Table 3: Environmental indicators by NOVA class of consumption according to %UPF quintiles (in g)[1], (INCA 3, n=2,121)[1]**

| | | Q1 | Q2 | Q3 | Q4 | Q5 |
|---|---|---|---|---|---|---|
| Climate change | NOVA1 | 4.23 (4.02-4.44) | 3.95 (3.74-4.15) | 3.55 (3.34-3.76) | 3.66 (3.45-3.87) | 3.25 (3.04-3.46) |
| (GHGe) | NOVA2 | 0.20 (0.19-0.21) | 0.18 (0.16-0.19) | 0.16 (0.15-0.18) | 0.15 (0.14-0.16) | 0.14 (0.12-0.15) |
| (kg CO2 eq) | NOVA3 | 0.65 (0.62-0.68) | 0.58 (0.55-0.60) | 0.57 (0.54-0.60) | 0.50 (0.47-0.53) | 0.43 (0.40-0.46) |
| | UPF (NOVA4) | 0.87 (0.79-0.94) | 1.16 (1.08-1.23) | 1.32 (1.24-1.40) | 1.63 (1.55-1.71) | 2.02 (1.94-2.09) |
| | *% from UPF* | 15% | 20% | 24% | 27% | 35% |
| Water use | NOVA1 | 5.64 (5.41-5.87) | 4.55 (4.32-4.78) | 4.24 (4.02-4.47) | 3.61 (3.38-3.84) | 3.15 (2.92-3.37) |
| ($m^3$ world eq) | NOVA2 | 0.24 (0.22-0.27) | 0.19 (0.17-0.22) | 0.16 (0.14-0.19) | 0.16 (0.13-0.18) | 0.14 (0.12-0.17) |
| | NOVA3 | 0.64 (0.60-0.67) | 0.57 (0.53-0.61) | 0.57 (0.54-0.61) | 0.47 (0.43-0.50) | 0.42 (0.39-0.46) |
| | UPF (NOVA4) | 0.83 (0.74-0.92) | 1.27 (1.18-1.37) | 1.34 (1.25-1.44) | 1.69 (1.60-1.78) | 2.37 (2.27-2.46) |
| | *% from UPF* | *11%* | *19%* | *21%* | *28%* | *39%* |
| Land use | NOVA1 | 203.09 (190.29-215.89) | 191.19 (178.56-203.82) | 173.56 (160.88-186.25) | 182.75 (170.05-195.45) | 170.65 (157.95-183.34) |
| (pt) | NOVA2 | 9.32 (8.78-9.87) | 8.69 (8.16-9.23) | 7.90 (7.36-8.44) | 7.42 (6.88-7.96) | 6.91 (6.37-7.44) |
| | NOVA3 | 34.82 (33.28-36.37) | 31.07 (29.55-32.58) | 32.45 (30.93-33.97) | 26.74 (25.22-28.27) | 23.68 (22.14-25.21) |
| | UPF (NOVA4) | 35.21 (31.32-39.11) | 44.92 (41.08-48.76) | 51.67 (47.81-55.53) | 63.34 (59.48-67.20) | 79.61 (75.75-83.47) |
| | *% from UPF* | *12%* | *16%* | *19%* | *23%* | *28%* |
| Energy demand | NOVA1 | 44.17 (43.10-45.24) | 38.89 (37.84-39.94) | 35.36 (34.30-36.41) | 32.55 (31.49-33.61) | 27.40 (26.35-28.46) |
| (MJ) | NOVA2 | 1.21 (1.15-1.28) | 1.09 (1.02-1.15) | 0.98 (0.92-1.05) | 0.92 (0.85-0.99) | 0.82 (0.76-0.89) |
| | NOVA3 | 9.61 (9.18-10.05) | 8.72 (8.29-9.14) | 8.68 (8.25-9.11) | 7.09 (6.66-7.52) | 6.36 (5.93-6.79) |
| | UPF (NOVA4) | 8.55 (7.91-9.18) | 12.51 (11.89-13.14) | 15.10 (14.47-15.73) | 18.91 (18.28-19.54) | 24.94 (24.31-25.58) |
| | *% from UPF* | *13%* | *20%* | *25%* | *32%* | *42%* |

Abbreviations: GHGe, greenhouse gas emissions

[1]Values are energy-adjusted means (95%CI), all P-values for linear trend (estimated using linear contrast) < 0.001

**Supplemental Table 4: Daily diet-related environmental indicators according to %UPF quintiles (in g), Sensitivity analyses (INCA 3, N=2,121)**

| Adjusted for dietary energy intake and %NOVA1 | Q1 | Q2 | Q3 | Q4 | Q5 | P-trend[2] |
|---|---|---|---|---|---|---|
| Climate change (GHGe) (kg $CO_2$ eq) | 5.23 (0.14) | 5.47 (0.12) | 5.54 (0.11) | 6.15 (0.11) | 6.88 (0.16) | <.0001 |
| Water use ($m^3$ world eq) | 6.56 (0.15) | 6.17 (0.13) | 6.27 (0.12) | 6.17 (0.13) | 7.24 (0.19) | 0.04 |
| Land use (pt) | 251.02 (8.19) | 257.88 (6.99) | 261.51 (6.52) | 287.50 (6.71) | 322.90 (9.86) | <.0001 |
| Energy demand (MJ) | 58.82 (0.75) | 58.65 (0.64) | 59.71 (0.59) | 60.80 (0.61) | 66.27 (0.90) | <.0001 |
| Acidification (mol H+ eq) | 0.07 (0.00) | 0.07 (0.00) | 0.07 (0.00) | 0.07 (0.00) | 0.08 (0.00) | <.0001 |
| Resource use, minerals and metals (kg Sb eq) | 10.17 (0.16) | 9.91 (0.13) | 9.42 (0.12) | 9.29 (0.13) | 9.48 (0.19) | 0.003 |
| Eutrophication, freshwater (kg P eq) | 1.00 (0.03) | 0.99 (0.02) | 0.96 (0.02) | 0.97 (0.02) | 1.10 (0.03) | 0.11 |
| Eutrophication, marine (kg N eq) | 21.27 (0.47) | 21.79 (0.40) | 21.97 (0.38) | 22.40 (0.39) | 24.48 (0.57) | 0.001 |
| Eutrophication, terrestrial (mol N eq) | 0.26 (0.01) | 0.26 (0.01) | 0.27 (0.01) | 0.29 (0.01) | 0.32 (0.01) | <.0001 |
| Photochemical ozone formation (kg NMVOC eq) | 16.05 (0.44) | 16.04 (0.37) | 15.56 (0.35) | 16.69 (0.36) | 18.02 (0.53) | 0.02 |
| Ozone depletion (kg CFC-11eq) | 0.47 (0.05) | 0.51 (0.04) | 0.47 (0.04) | 0.67 (0.04) | 0.63 (0.06) | 0.03 |
| Particulate matter (disease incidence) | 0.48 (0.01) | 0.49 (0.01) | 0.48 (0.01) | 0.52 (0.01) | 0.57 (0.01) | 0.00002 |
| Ionising radiation (kBq U235 eq) | 1.31 (0.02) | 1.31 (0.02) | 1.36 (0.01) | 1.38 (0.02) | 1.48 (0.02) | <.0001 |
| EF score[3] | 0.66 (0.01) | 0.66 (0.01) | 0.66 (0.01) | 0.70 (0.01) | 0.77 (0.01) | <.0001 |

Abbreviation: EF, ecological footprint; GHGe, greenhouse gas emissions. Units are as follows: kg CO2 eq, carbon dioxide equivalent; m3 world eq, water use in cubic meters of water; land use is estimated as loss of soil organic matter content in kilograms of carbon deficit (kg C deficit) dimensionless and expressed as Points (Pt); MJ, megajoule; mol H+ eq, equivalent of moles hydron; kg Sb eq, equivalent of kilograms of antimony; kg P eq, equivalent of kilograms of phosphorus, kg N eq, equivalent of kilograms of nitrogen; mol N eq, equivalent of moles of nitrogen; kg NMVOC eq, equivalent of kilograms of non-methane volatile organic compounds; kg CFC-11eq, equivalent of kilograms of trichloroflurومethane (Freon-11); Emission of particulate matter in change in mortality due to particulate matter emissions; kg U235 eq, equivalent of kilobecquerels of Uranium 235

[1]Values are means (standard error of the mean) adjusted for energy intake and %NOVA1

[2]P-value for linear trend is estimated using contrast

[3]For the EF score, the higher it is, the more impactful it is

**Supplemental Table 5: Daily diet-related environmental indicators according to %UPF quartiles (in kcal), (INCA 3, N=2,121)**

| **Crude model[1]** | **Q1** | **Q2** | **Q3** | **Q4** | **P for trend[2]** |
|---|---|---|---|---|---|
| Climate change (GHGe ) (kg CO2 eq) | 5.30 (2.30) | 5.93 (2.47) | 5.93 (2.47) | 6.53 (3.88) | <.0001 |
| Water use ($m^3$ world eq) | 6.94 (3.18) | 6.35 (2.83) | 6.68 (2.92) | 6.19 (2.77) | 0.0007 |
| Land use (pt) | 255.80 (136.39) | 287.14 (140.61) | 279.19 (148.44) | 297.51 (205.96) | 0.0001 |
| Energy demand (MJ) | 58.36 (17.21) | 61.58 (17.69) | 62.21 (17.87) | 64.16 (25.33) | <.0001 |
| Acidification (mol H+ eq) | 0.065 (0.028) | 0.072 (0.031) | 0.070 (0.031) | 0.075 (0.046) | <.0001 |
| Resource use, minerals and metals (kg Sb eq) | 9.43 (3.25) | 9.85 (3.53) | 9.98 (3.23) | 9.92 (4.40) | 0.02 |
| Eutrophication, freshwater (kg P eq) | 1.00 (0.56) | 0.99 (0.43) | 1.04 (0.55) | 1.02 (0.51) | 0.26 |
| Eutrophication, marine (kg N eq) | 21.04 (8.15) | 23.52 (11.02) | 22.94 (9.44) | 23.37 (11.89) | 0.0012 |
| Eutrophication, terrestrial (mol N eq) | 0.26 (0.12) | 0.29 (0.13) | 0.28 (0.13) | 0.30 (0.19) | <.0001 |
| Photochemical ozone formation (kg NMVOC eq) | 15.85 (7.41) | 17.05 (7.85) | 16.63 (7.72) | 17.09 (9.89) | 0.04 |
| Ozone depletion (kg CFC-11eq) | 0.49 (0.42) | 0.57 (0.76) | 0.52 (0.29) | 0.64 (1.59) | 0.0201 |
| Particulate matter (disease incidence) | 0.47 (0.19) | 0.53 (0.21) | 0.51 (0.21) | 0.55 (0.32) | <.0001 |
| Ionising radiation (kBq U235 eq) | 1.30 (0.42) | 1.39 (0.42) | 1.40 (0.42) | 1.46 (0.61) | <.0001 |
| EF score | 0.65 (0.23) | 0.71 (0.25) | 0.70 (0.25) | 0.74 (0.37) | <.0001 |
| **Adjustment for EI[3]** | **Q1** | **Q2** | **Q3** | **Q4** | **P for trend[2]** |
| Climate change (GHGe) (kg CO2 eq) | 5.59 (0.10) | 5.87 (0.10) | 5.65 (0.10) | 6.18 (0.10) | <.0001 |
| Water use (m3 world eq) | 7.18 (0.11) | 6.29 (0.11) | 6.44 (0.11) | 5.88 (0.11) | 0.0007 |
| Land use (pt) | 269.72 (5.88) | 283.98 (5.85) | 265.41 (5.88) | 279.99 (5.88) | 0.0001 |
| Resource use, fossils (MJ) | 60.90 (0.55) | 61.01 (0.54) | 59.70 (0.55) | 60.97 (0.55) | <.0001 |
| Acidification (mol H+ eq) | 0.07 (0.00) | 0.07 (0.00) | 0.07 (0.00) | 0.07 (0.00) | <.0001 |
| Resource use, minerals and metals (kg Sb eq) | 9.85 (0.11) | 9.76 (0.11) | 9.56 (0.11) | 9.40 (0.11) | 0.02 |
| Eutrophication, freshwater (kg P eq) | 1.04 (0.02) | 0.98 (0.02) | 1.00 (0.02) | 0.97 (0.02) | 0.26 |
| Eutrophication, marine (kg N eq) | 22.14 (0.34) | 23.27 (0.34) | 21.84 (0.34) | 21.97 (0.34) | 0.0012 |
| Eutrophication, terrestrial (mol N eq) | 0.27 (0.01) | 0.29 (0.01) | 0.27 (0.01) | 0.29 (0.01) | <.0001 |
| Photochemical ozone formation (kg NMVOC eq) | 16.51 (0.31) | 16.90 (0.31) | 15.98 (0.31) | 16.26 (0.31) | 0.04 |
| Ozone depletion (kg CFC-11eq) | 0.51 (0.04) | 0.56 (0.04) | 0.49 (0.04) | 0.61 (0.04) | 0.02 |
| Particulate matter (disease incidence) | 0.50 (0.01) | 0.52 (0.01) | 0.49 (0.01) | 0.52 (0.01) | <.0001 |
| Ionising radiation (kBq U235 eq) | 1.36 (0.01) | 1.38 (0.01) | 1.34 (0.01) | 1.38 (0.01) | <.0001 |
| EF score | 0.68 (0.01) | 0.70 (0.01) | 0.67 (0.01) | 0.70 (0.01) | <.0001 |

Abbreviation: EF, ecological footprint; GHGe, greenhouse gas emissions. Units are as follows: kg CO2 eq, carbon dioxide equivalent; m3 world eq, water use in cubic meters of water; land use is estimated as loss of soil organic matter content in kilograms of carbon deficit (kg C deficit) dimensionless and expressed as Points (Pt); MJ, megajoule; mol H+ eq, equivalent of moles hydron; kg Sb eq, equivalent of kilograms of antimony; kg P eq, equivalent of kilograms of phosphorus, kg N eq, equivalent of kilograms of nitrogen; mol N eq, equivalent of moles of nitrogen; kg NMVOC eq, equivalent of kilograms of non-methane volatile organic compounds; kg CFC-11eq, equivalent of kilograms of trichlorofluromethane (Freon-11); Emission of particulate matter in change in mortality due to particulate matter emissions; kg U235 eq, equivalent of kilobecquerels of Uranium 235

[1]Values are means (SD)

[2]P-value for linear trend is estimated using linear contrast

[3]Values are means (standard errors of the means)

**Supplemental Figure 1: contribution of UPF according to food groups[1]**

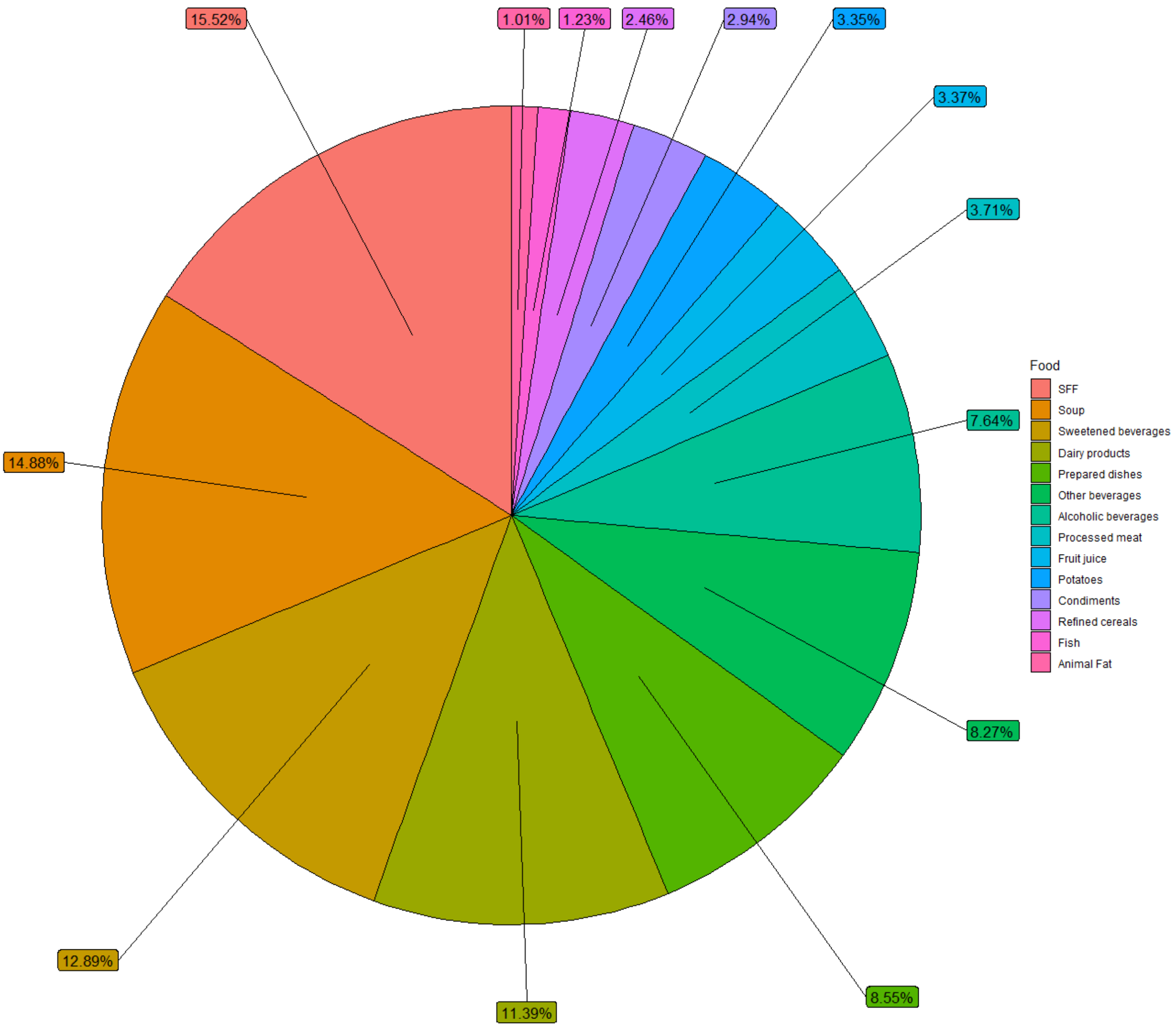


Abbreviation: SFF: sweet and fat food
[1]Food groups whose contribution to less than 1% to ultra-processed food consumption were removed (legumes, nuts, poultry, eggs, meat, vegetables, wholegrains, fruits, vegetable oils)

**Supplemental Figure 2: Contribution of food groups to environmental indicators[1]**

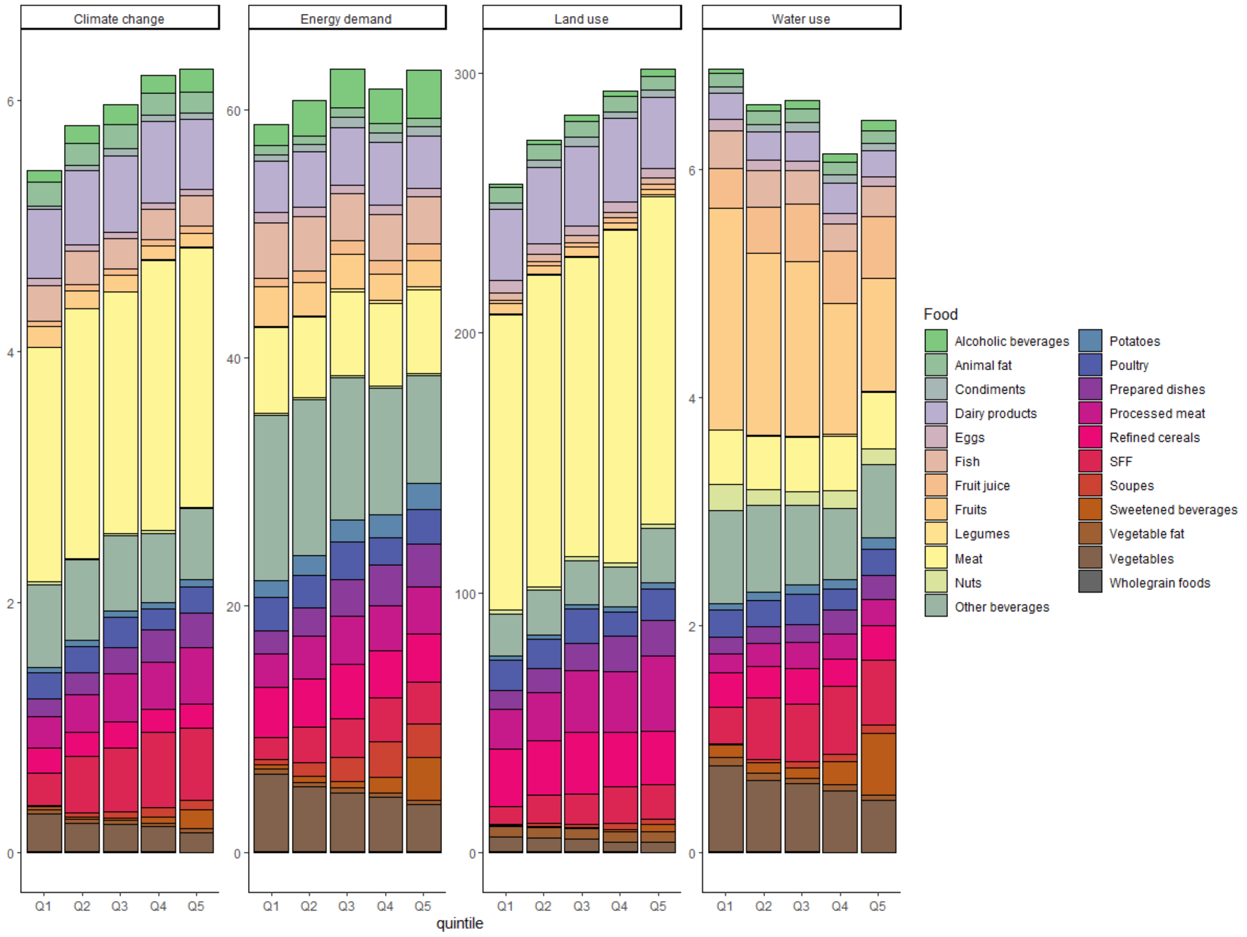


Abbreviations: SFF, sweet and fat food
[1]Climate change (Greenhouse gas emissions), energy demand, land use and water use are expressed in kg CO2eq, $m^3$ world eq, pt, MJ, respectively
[2]Values are mean of consumption (g/d)
